# Modeling trajectories of routine blood tests as dynamic biomarkers for outcome in spinal cord injury

**DOI:** 10.1101/2025.01.20.25320728

**Authors:** Marzieh Mussavi Rizi, Daniel Fernandez, John LK Kramer, Rajiv Saigal, Anthony M. DiGiorgio, Michael S. Beattie, Adam R Ferguson, Nikos Kyritsis, Abel Torres-Espin, TRACK-SCI investigators

## Abstract

**Importance:** Early outcome prediction after acute traumatic spinal cord injury (SCI) is challenging due to pathological complexities and population heterogeneity. Routinely collected data during standard medical practice, such as laboratory analytics, can be a surrogate of underlying pathophysiological processes and used as a biomarker. We hypothesized that distinct temporal trends of blood analytics could be modeled after SCI and that those would predict distinct outcome parameters.

**Objective:** To test the hypothesis and develop machine learning models for predicting SCI outcomes.

**Design:** We developed and validated the models using retrospective data from the MIMIC-III and MIMIC-IV datasets and the prospective TRACK-SCI study, covering the period from 2001 to 2020.

**Setting:** Multi-center, involving data obtained from intensive care units across several different hospital settings in the United States.

**Participants:** Patients 15 years and older with traumatic SCI or vertebral fractures, admitted to emergency facilities, were included, resulting in a final cohort of 2,615 patients for modeling.

**Exposure(s):** NA

**Main Outcome(s) and Measure(s):** Primary outcomes included in-hospital mortality, occurrence of SCI and vertebral fracture in spine trauma patients, and SCI severity measured by the ASIA Impairment Scale. Blood biomarker level trajectory memberships served as predictors.

**Results:** Our study analyzed 2,752 patients, comprising 2,615 from the MIMIC dataset and 137 from the TRACK-SCI study. We identified multiple trajectory classes for 20 common blood markers that serve as dynamic predictors in machine learning classifiers. The in-hospital mortality model achieved an area under the Precision-Recall curve (PR-AUC) of 0.92 in the training set by leveraging trajectory data and baseline covariates from as early as day one post-injury. For SCI severity, the models distinguished between complete and incomplete motor outcomes with a PR-AUC of 0.78. The trajectory-based models showed significant improvement over traditional severity scores, such as Simplified Acute Physiology Score (SAPS) II, especially when combined with demographic information.

**Conclusions and Relevance:** Real-world routinely obtained blood test data can be used to model dynamic changes after SCI with prediction validity for patient outcomes. This work establishes the basis for further development of dynamic biomarker data for outcome prediction in neurotrauma and other neurological conditions.

**Key Points:** *Question:* Can dynamic changes of routinely collected acute blood test data serve as biomarkers to predict outcomes in patients with traumatic spinal cord injury (SCI)?

*Findings:* In this study using data from the MIMIC and TRACK-SCI datasets, we developed machine learning models that categorize patients into distinct groups based on the temporal and non-linear dynamics of blood biomarkers. These models effectively predicted in-hospital mortality and SCI severity, indicating significant predictive utility from as early as the first day of hospitalization.

*Meaning:* The application of dynamic machine learning models to blood test data has potential to significantly predict the prognosis and enhance management of traumatic spinal cord injury in clinical settings.

## Introduction

Traumatic spinal cord injury (SCI) presents a global health challenge, with an estimated 930,000 new cases and a prevalence of 17 million worldwide in 2016, causing significant personal and economic burdens.^1,2^ The complexity of SCI, characterized by variable clinical presentations and recovery trajectories, complicates acute diagnosis and prognosis.^3–8^ Early neurological assessment of SCI is the main tool for characterizing the injury; however, it is limited by dependence on patient responsiveness and the presence of comorbid injuries.^9^ Other objective measures such as MRI,^10,11^ physiological time-series,^12–14^ and fluid biomarkers^15–21^ are being investigated, though their accessibility across medical settings might be limited.^5^

There is a growing interest in utilizing routinely collected in-hospital data, such as blood laboratory values, as biomarkers.^15,16^ Spinal cord damage triggers pathophysiological cascades of measurable events in the blood,^20,22^ and hematologic abnormalities after SCI correlate with injury severity and neurological outcome.^22,23^ However, the correlation between a specific marker and critical aspects of the injury—such as its severity, location, and the patient’s neurological recovery—is poor.^15,16,21^ Moreover, any marker may be influenced by factors other than SCI, including demographics, concomitant injuries, and other comorbidities. The inherent dynamic and non-linear progression of SCI, coupled with the asynchronous evolution of routine in-hospital data, further complicate this-

We hypothesize that routine blood biomarkers and their changes over time provide valuable predictive information for SCI-related outcomes. Capturing the evolution of blood biomarkers during a patient’s hospital stay and their relationship with SCI over time represents a novel perspective that has not been widely explored. To test this hypothesis, we propose machine learning models to predict SCI outcomes using longitudinal data from routine blood tests, leveraging data from two sources: the electronic health records (EHR) of the Medical Information Mart for Intensive Care (MIMIC^24^) and the Transforming Research and Clinical Knowledge in Spinal Cord Injury (TRACK-SCI^25^) study.

Our model captures the multidimensional, nonlinear, and temporal changes in multiple blood markers over time, rather than relying on single measurements at static time points. We first modeled blood biomarker trajectories using longitudinal finite mixture models, identifying distinct patient subgroups with unique pathophysiological profiles. The membership of these trajectory-based subgroups was then used as dynamic predictors in machine learning classifiers to predict in-hospital mortality and SCI severity.

Our findings demonstrate that longitudinal changes in blood lab values can serve as effective dynamic biomarkers for outcomes after SCI. In practice, this approach could potentially provide real-time outcome predictions based on information available at each time point, enabling earlier identification of SCI patients at higher risk of adverse outcomes and allowing clinicians to intervene proactively.

## Methods

### Study Population

Two different epochs of the MIMIC dataset (MIMIC-III and the MIMIC-IV) were accessed under a data use agreement through the PhysioNet project.^26^ We identified patients 15 years and older with traumatic SCI or spine trauma (vertebral fracture), based on ICD9/ICD10 diagnostic codes for emergency admissions.^27^ For each patient, we selected their first hospital admission that matched these criteria. We harmonized patient demographic data into one dataset and categorized patients into three groups based on their diagnoses: SCI with vertebral fracture (SCI Fracture), SCI without vertebral fracture (SCI no Fracture), and spine trauma without SCI (Spine Trauma).To classify trajectories in new patients, we used de-identified data from 137 patients enrolled in the TRACK-SCI study (details in eMethods, Supplement 1) obtained through a collaboration agreement. Data collection and extraction protocols for the TRACK-SCI study were approved by the Institutional Research Board (IRB) at the University of California, San Francisco.

### Modeling data

#### Laboratory values

We extracted laboratory values during hospital stays and calculated the time from admission to laboratory sample collection. Among 413 distinct laboratory markers (157 hematology, 161 chemistry, 15 blood gases; eTable 1 in Supplement 1), we focused on the 20 most common, measured in 90–98% of patients, as the modeling set. This set was pre-processed for outliers and limited to data within 21 days of admission to address the drop in available patient data over time (eFigures 1-3 in Supplement 1).

#### Demographics and Hospital stay characteristics

Demographics included age, gender, ethnicity, and insurance type. Stay details included the length of stay (in days), number of ICD diagnostics, admission and discharge locations, and admission type. The analysis was restricted to emergency admissions.

#### Validation data

Laboratory names were harmonized to align TRACK-SCI with MIMIC markers, dynamic ranges were compared to ensure consistent scaling, and TRACK-SCI lab values underwent the same preprocessing as MIMIC. In TRACK-SCI, International Standards for Neurological Classification of Spinal Cord Injury (ISNCSCI) exams are performed multiple times during hospitalization. We used the latest exam available at hospital discharge to calculate the American Spinal Injury Association Impairment Severity (AIS) ^28^ grade, a 5-point scale (A to E) measuring neurological impairment after SCI, as the outcome (further details on data processing in eMethods of Supplement 1).

### Multi-class trajectory modeling

Following recommendations,^29^ we first applied Latent Class Growth Analysis (LCGA), for each modeling set marker, determining the optimal number of classes (1-5) and evaluating linear and polynomial-growth trajectories over time. We implemented three link functions: a linear link for continuous Gaussian markers, and a quadratic I-spline with three knots or Beta density function for other continuous markers. First, LCGA models were developed in R using the lcmm package without random effects.^30^ Model selection was based on the Bayes Information Criterion (BIC)^31^, Integrated Complete-data Likelihood (ICL)^29^, and the Average Posterior Probability of Assignment (APPA)^32^, prioritizing lower BIC and ICL values and an APPA above 0.7. Next, we refined LCGA results by using Growth Mixture Models (GMMs) with random effects and employing natural splines to allow for non-linear time trajectory modeling. Based on the GMM, posterior probabilities of assignment (PPA) were estimated and then used as predictors in an Elastic Net classification model. ^33^Multi-class trajectory modeling and model selection were performed on MIMIC data, with the selected models validated using the TRACK-SCI dataset (see eMethods, Supplement 1 for details on model selection).

### Machine learning classifiers and outcomes

Our study designed three predictive modeling experiments using Elastic Net^33^ models (“glmnet” model in the R caret package)^34^ to predict in-hospital mortality (Experiment I), SCI occurrence in spine trauma patients (Experiment II), and SCI severity based on the AIS grade in the TRACK-SCI cohort (Experiment III), with data cutoffs at 1, 3, 7, 14, and 21-days post-injury. Predictors included class trajectory probabilities for each marker, summary statistics of marker values, and demographic variables at admission: age, gender, ethnicity, and insurance type. Model training considered these predictor sets both individually and in combination. Each experiment type and cutoff combination was run 25 times with varying random seeds, using a fixed seed across cutoffs for comparability. We used an 80/20 training/testing split, applied down-sampling during training to balance class representation, and performed three-fold cross-validation to fine-tune hyperparameters. Model performance was evaluated using the Precision-Recall curve (PR-AUC) and ROC curve (ROC-AUC), calculated with the pROC R package. ^22^

Elastic Net is a regularized regression method that performs feature selection by shrinking coefficients, making it effective for handling highly correlated data. We applied it to assess the importance of variables across different predictor sets for each experiment.

We benchmarked model performance in Experiments I and II against the regressions on Simplified Acute Physiology Score (SAPS) II.^35^ This score measures disease severity and is widely used in ICUs (see eMethods, Supplement 1 for details on SAPS II).

### Statistics

Differences between trajectory groups and patient characteristics (age, gender, ethnicity, cohort group, length of hospital stay, whether the patient died in hospital, and the number of ICD diagnostics), were analyzed using ANOVA or t-test for continuous variables and Fisher exact test for categorical variables. P-values were adjusted for false discovery rate by the Benjamini-Hochberg method, and q-values were reported. The level of significance was set at q < 0.05. A Friedman sum of rank test was used to test for the null hypothesis of no differences between machine learning performance in Experiment I-III between feature lists at each time window cutoff or in each feature list over time.

## Results

### Patient cohort for routine blood trajectory modeling

A total of 1194 and 4529 unique patients were identified from MIMIC-III and MIMIC-IV, respectively. After filtering, a final cohort of 2615 patients with processed laboratory marker data were included for modeling (Figure 1).

**Figure 1.**
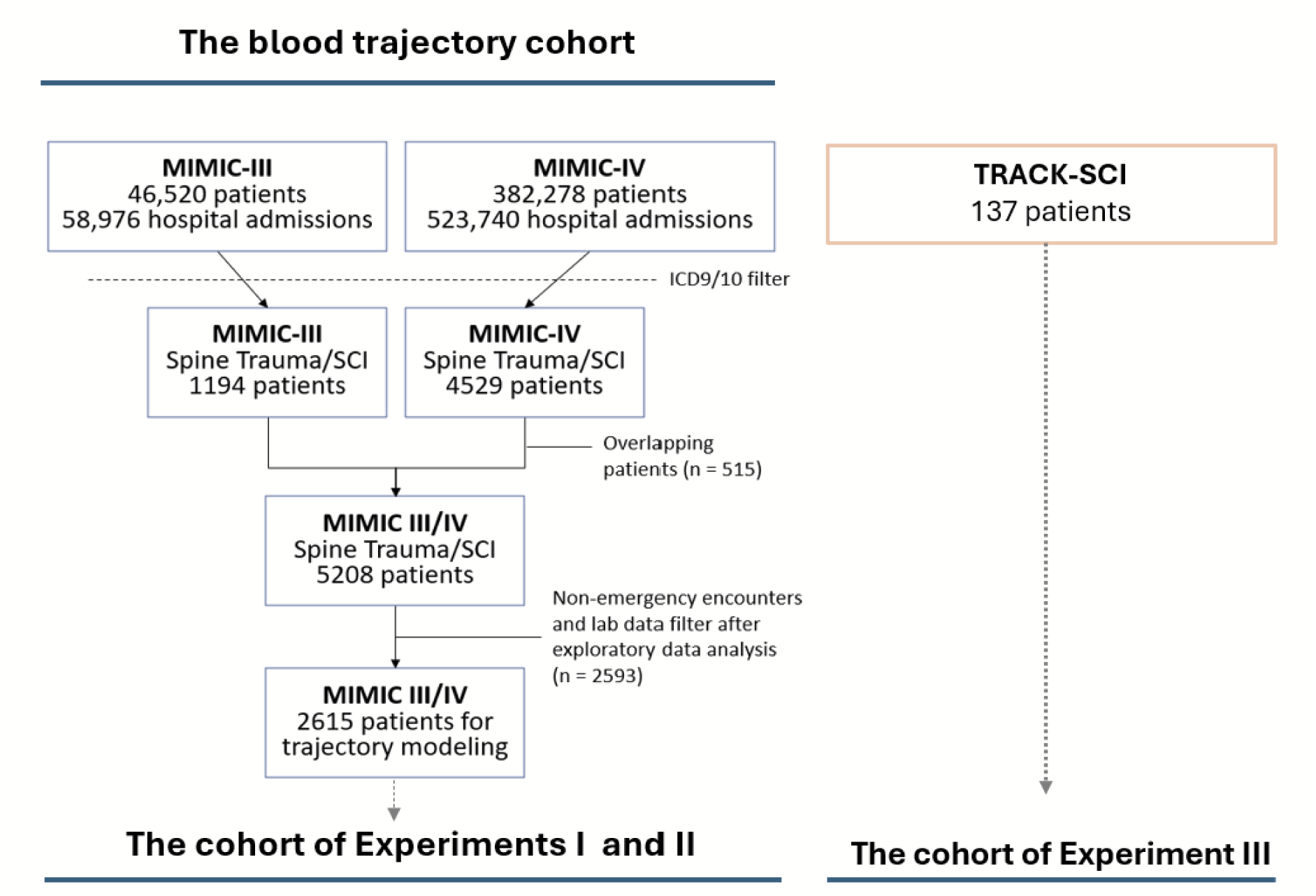
Flow diagram of cohort build. Patients from MIMIC-III/IV were first filtered based on their ICD9 and ICD10 diagnostic codes. Then, potential overlapping patients were filtered from MIMIC-IV. After laboratory analyte data extraction and data cleaning, patients with less than 3 measures for any of the 20 most common analytes were excluded. The data from a total of 2615 patients from both MIMIC databases were used for trajectory modeling as well as Experiments I and II. An additional 137 patients from the TRACK-SCI dataset were used as a validation set in Experiment III to predict SCI severity based on AIS grade.

Table 1 shows demographic variables for the selected cohorts from MIMIC datasets (SCI Fracture, SCI noFracture, and Spine Trauma). There were statistical differences in age, gender, insurance type, ethnicity, and dataset (i.e., MIMIC III or IV) across the three groups. The SCI Fracture cohort was younger and with higher proportions of males than the Spine Trauma patients. A higher proportion of patients in the SCI Fracture group came from the MIMIC-III database (60%), while in the SCI noFracture and Spine Trauma groups, ∼60% of patients came from the MIMIC-IV database. Table 2 shows the hospital stay characteristics for MIMIC. We observed statistical differences in length of stay and discharge location. On average, the SCI Fracture patients stayed in hospital 3 and 4 more days than SCI noFracture and the Spine Trauma groups, respectivley. SCI Fracture had a higher mortality rate (11%), and a higher proportion of patients were discharged to rehabilitation (54%) than the other two groups.

**Table 1.** Demographics for the MIMIC cohorts.

| *Characteristic* | **SCI**<br>**Fracture**<br>**N = 382<sup>1</sup>** | **SCI**<br>**noFracture**<br>**N = 125<sup>1</sup>** | **Spine**<br>**Trauma**<br>**N = 2,108<sup>1</sup>** | **p-**<br>**value<sup>2</sup>** | **q-**<br>**value<sup>3</sup>** |
| --- | --- | --- | --- | --- | --- |
| *Age* | 55 (39, 71) | 57 (44, 72) | 65 (44, 81) | <0.001 | **<0.001** |
| *Gender* |  |  |  | <0.001 | **<0.001** |
| *F* | 101 (26%) | 37 (30%) | 924 (44%) |  |  |
| *M* | 281 (74%) | 88 (70%) | 1,184 (56%) |  |  |
| *Insurance* |  |  |  | 0.011 | **0.011** |
| *Medicaid* | 37 (9.7%) | 13 (10%) | 154 (7.3%) |  |  |
| *Medicare* | 118 (31%) | 48 (38%) | 865 (41%) |  |  |
| *Other* | 221 (58%) | 62 (50%) | 1,064 (50%) |  |  |
| *Other Government* | 6 (1.6%) | 2 (1.6%) | 25 (1.2%) |  |  |
| *Ethnicity* |  |  |  | <0.001 | **<0.001** |
| *ASIAN* | 6 (1.9%) | 0 (0%) | 43 (2.3%) |  |  |
| *BLACK/AFRICAN*<br>*AMERICAN* | 14 (4.4%) | 23 (20%) | 88 (4.8%) |  |  |
| *HISPANIC/LATINO* | 14 (4.4%) | 9 (7.9%) | 77 (4.2%) |  |  |
| *MULTI RACE/ETHNICITY* | 1 (0.3%) | 0 (0%) | 5 (0.3%) |  |  |
| *OTHER* | 17 (5.4%) | 5 (4.4%) | 84 (4.6%) |  |  |
| *WHITE* | 264 (84%) | 77 (68%) | 1,540 (84%) |  |  |
| *Unknown* | 66 | 11 | 271 |  |  |
| *Dataset* |  |  |  | <0.001 | **<0.001** |
| *MIMIC-III* | 231 (60%) | 49 (39%) | 826 (39%) |  |  |
| *MIMIC-IV* | 151 (40%) | 76 (61%) | 1,282 (61%) |  |  |

<sup>1</sup> Median (IQR); n (%)
<sup>2</sup> Kruskal-Wallis rank sum test; Fisher's Exact Test for Count Data with simulated p-value (based on 2000 replicates)
<sup>3</sup> False discovery rate correction for multiple testing

**Table 2.**
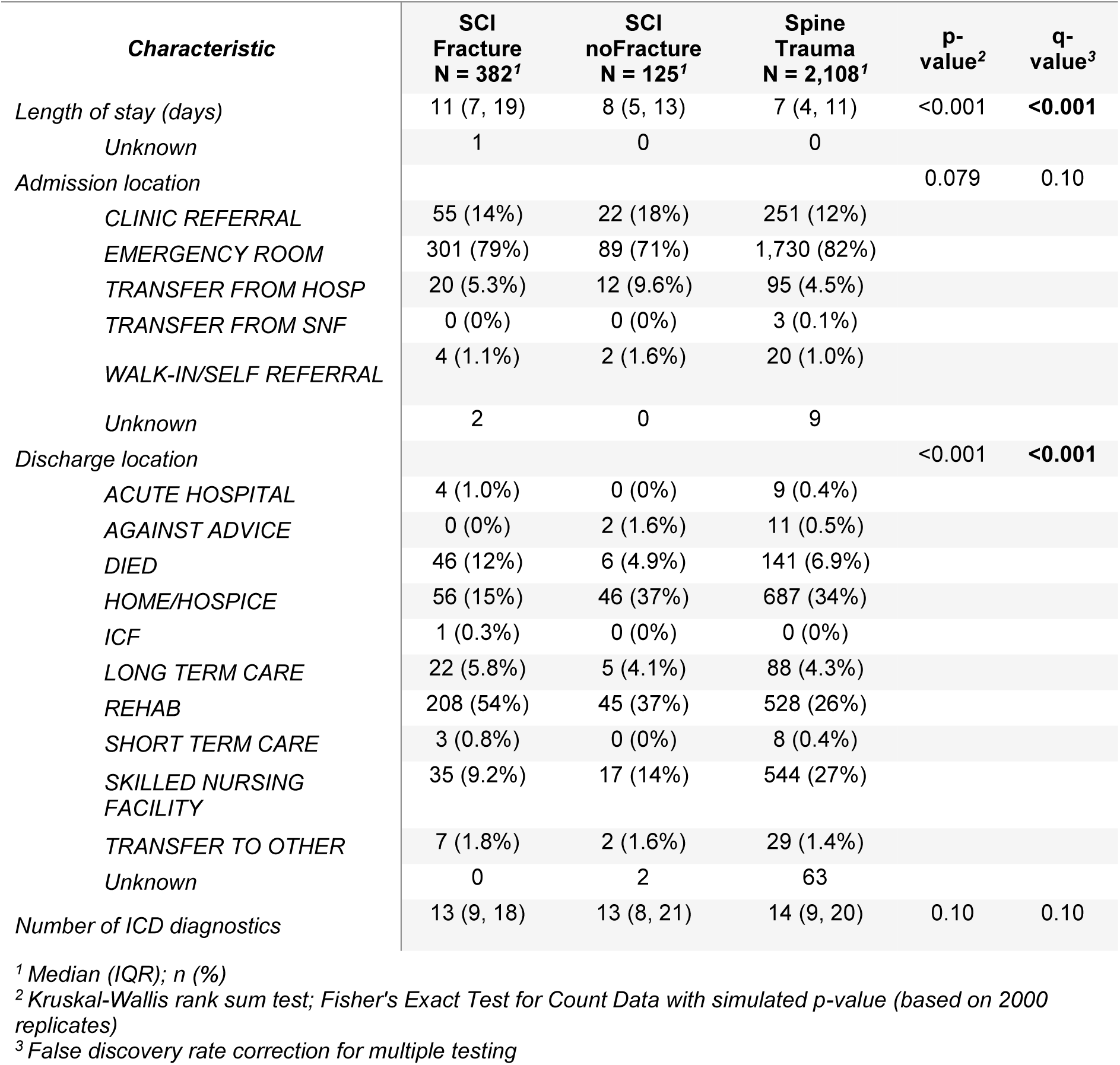
Hospital stay characteristics for the MIMIC cohorts.

### Routine blood trajectory models

Blood trajectories for the 20 most common markers for up to 21 days from hospital admission were modeled using longitudinal finite mixture models.^36^ To explore their use as dynamic biomarkers, we then set three dynamic prediction modeling experiments, using the latent trajectory memberships of each marker as predictors in a machine learning classifier (Fig. 2a-e).

**Figure 2.**
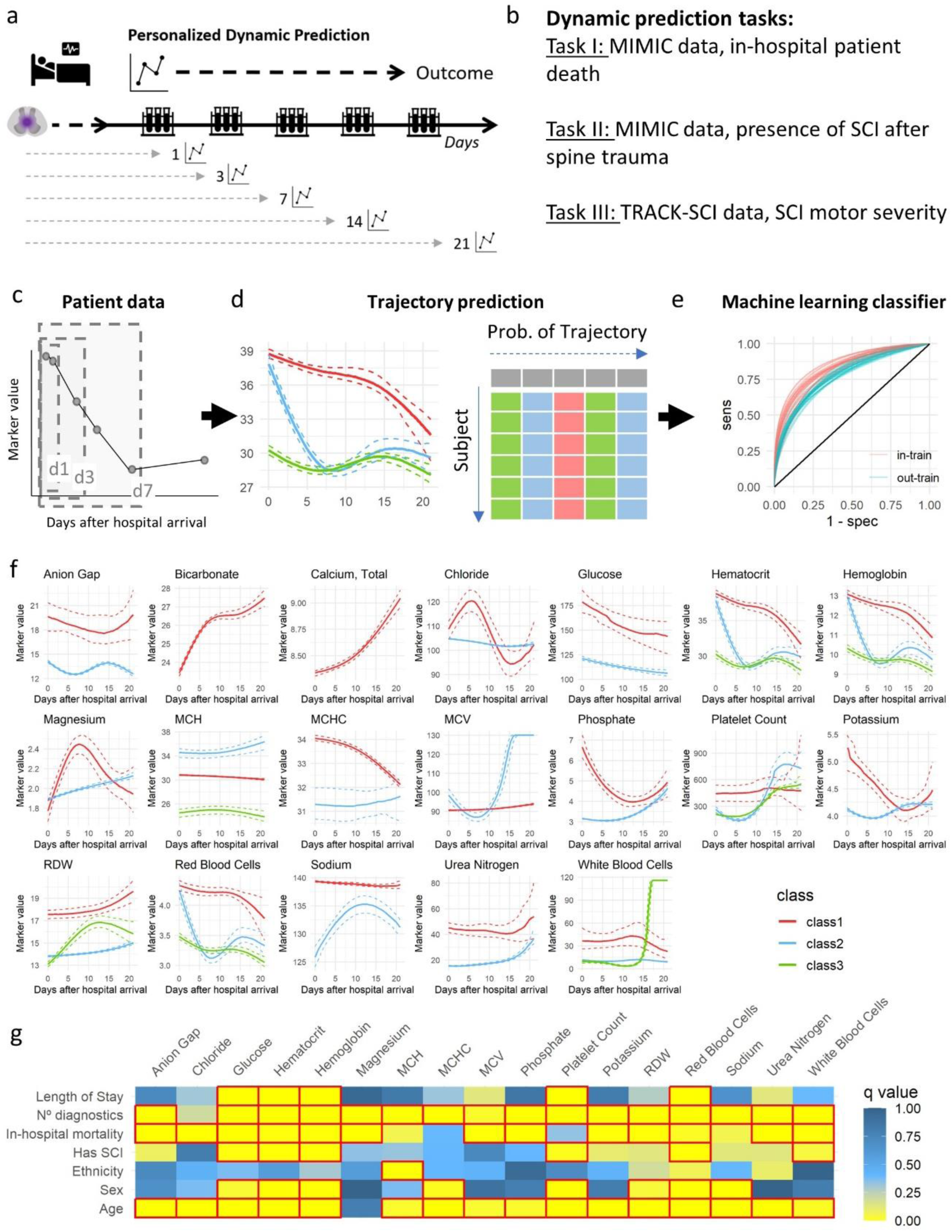
Dynamic prediction with blood trajectories. (a) Schematic of the dynamic prediction experiments simulating increased data availability over time after hospital arrival. (b) Three dynamic prediction tasks were set up to study the potential use of blood trajectories as biomarkers. (c) For each individual subject, laboratory data at different time windows is used to predict subject-specific trajectory probability of membership based on the trajectory models (d). This probability is then used as predictors in machine learning classifiers (e). (f)The mean trajectory for each one the classes for each analyte model are shown, together with the 95% CI. Note that although colored the same for visualization, these are univariate models and therefore classes might not be constituted by the same subjects across analytes. (g) Univariate summary of association of the different trajectory classes with demographics and other variables of interest. The data is shown as a heatmap of the q value (adjusted p value for multiple comparisons). Red boxes indicate a q value < 0.05.

Fig. 2f shows the predicted trajectories; the final selected models for each blood marker are presented in eTable 2 (Supplement 1). Creatinine models did not converge and were excluded. Except for bicarbonate and calcium, models with more than one class were the best fit. Anion gap, chloride, creatinine, magnesium, MCH, MCV, phosphate, platelet count, potassium, sodium, urea nitrogen, and white blood cells selected models had one dominant class containing above 90% of the subjects, often with a small secondary class. Biomarkers glucose, hematocrit, hemoglobin, MCHC, RDW, and red blood cells presented a more balanced distribution of subjects across classes. All markers except glucose were better modeled with non-linear time transformations, and 11 of 20 blood values were best modeled with non-Gaussian link functions.

Univariate analysis comparing class trajectories for each marker across different demographics and clinical characteristics is summarized in Fig. 2g and provided in more detail in eTables 3 to 11 in Supplement 1. For most blood biomarkers, significant differences were observed in age, in-hospital mortality rates, and the number of diagnostics across trajectory classes. Classes with fewer subjects were associated with higher in-hospital mortality rates and a higher number of diagnostics. For example, being in the Potassium Class 1 trajectory, characterized by higher marker levels early in hospitalization and a rapid decline to levels similar to Class 2 (Fig. 2f), is associated with an increased mortality rate.Length of stay and cohort group distribution also varied across different classes for several markers (eFigure 4 in Supplement 1).

### Blood trajectories as dynamic biomarkers for SCI diagnosis and prognosis

For Experiment I and II, models using only the trajectory list yielded high PR-AUC values for predictions made within the training sample across all time points, demonstrating strong performance even with data from just the first day post-admission (PR-AUC = 0.78 for in-hospital mortality, and 0.68 for SCI presence). Model performance increases over time and with the addition of summary statistics and demographic predictors (eFigures 8 in Supplement 1). A reduction in performance on the test sample was observed across all time points (Fig. 3a) as expected. However, it remained significantly above the non-information rate. Similar trends were observed for ROC-AUC (eFigures 8-14 in Supplement 1).

**Figure 3.**
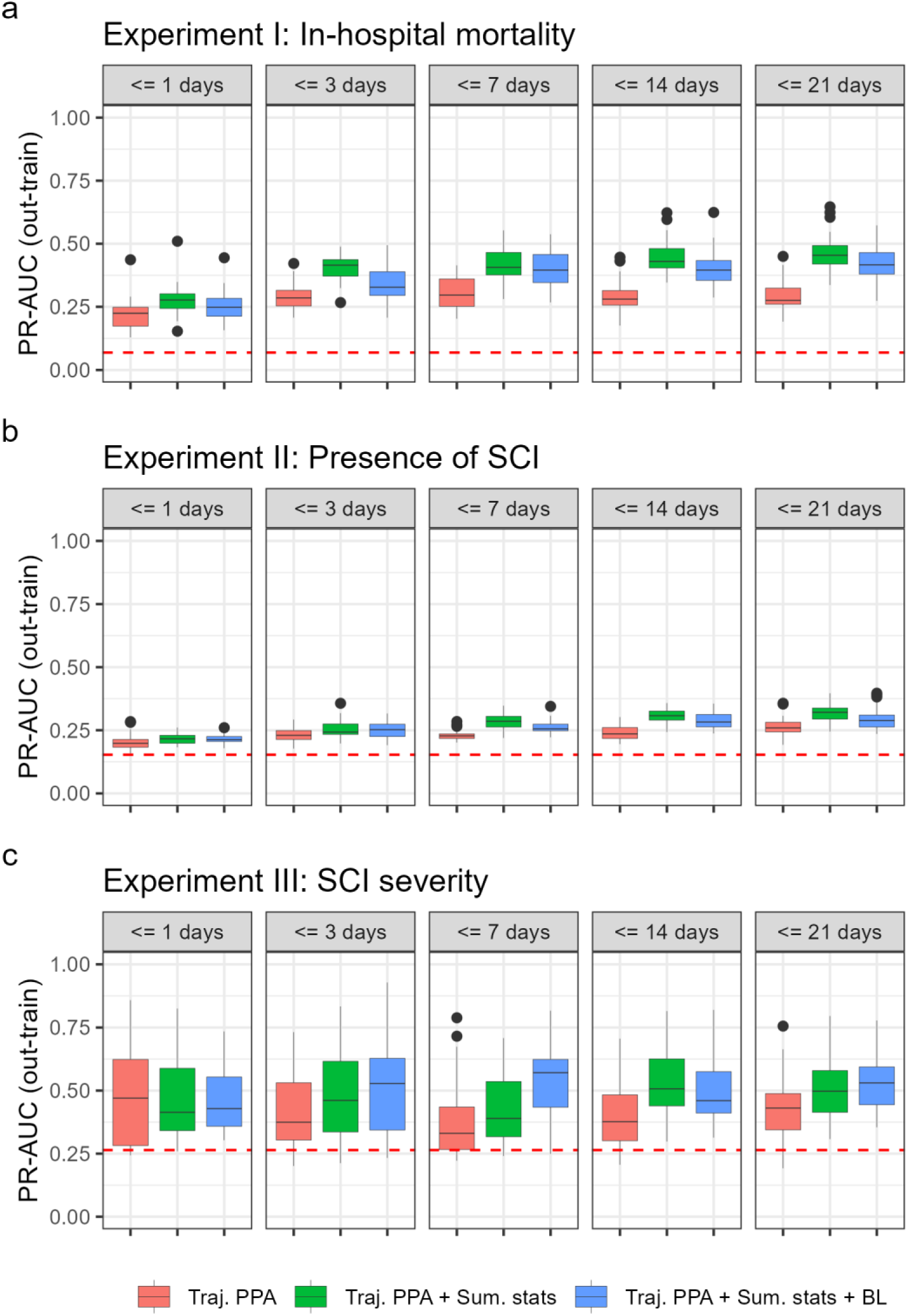
PR-AUC performance of dynamic predictions. (a) PR-AUC out-of-train sample performance of experiment I in-hospital mortality. (b) PR-AUC out-of-train sample performance of experiment II for detecting the presence of SCI after spine trauma. (c) PR-AUC out-of-train sample performance of experiment III on detecting SCI severity on the TRACK-SCI cohort, external to trajectory modeling. Dashed red lines represent the mean prevalence of the outcome of interest in each experiment. Three predictors’ lists are shown: Traj. PPA = posterior probability of class assignment only; + Sum. stats = addition of summary statistics of blood data; and + BL = addition of baseline predictors.

We benchmarked these models by comparing their performance to models using only SAPS II scores as predictors. Since, SAPS II scores were limited to ICU patients, the sample was restricted to ICU admissions. When compared to our models from Experiment I and Experiment II, models incorporating trajectories, summary statistics, and baseline covariates outperformed those using SAPS II alone (eFigures 12-13, Supplement 1).

Experiment III was set to classify whether SCI patients in the TRACK-SCI (not used during trajectory modeling) cohort presented motor complete (AIS A or B) vs. motor incomplete (AIS C, D or E) neurological deficits in its latest available in-hospital timepoint. Similar to Experiments I and III, PR-AUC for in-train sample was higher than out-train, reaching values as high as 0.98 (Fig. 3c). The addition of summary statistics and demographics increased PR-AUC performance, reaching 0.99 at up to 3 days and 0.98 up to 7 days of data.

We also identified the most important predictors for each model using the importance metric. Across the three experiments with different models, the blood biomarker posterior probability of assignment (PPA) consistently emerged among the most important variables. This consistency highlights their relevance to predicting the SCI outcome of interest. For example, the PPAs of white blood cell count, potassium, chloride, MCV and magnesium were among important predictors of in-hospital mortality (eFigures 15–17 in Supplement 1).

## Discussion

Secondary spinal cord damage triggers pathophysiological cascades of measurable events detectable in the blood,^20,22^ and hematologic abnormalities after SCI correlate with clinical metrics of injury severity.^22,23^ Leister et al. reported temporal changes in different blood metrics from a few hours to weeks to a year after SCI predicting whether patients will walk one year after injury.^21^ Furthermore, combining blood markers in static prognostic models with clinical characteristics improves prediction performance.^13,17,18^ We demonstrated that changes in routinely collected in-hospital blood test longitudinal data can be modeled and their trajectories used as dynamic biomarkers in machine learning for diagnosis and prognosis in SCI.. We also show that predictability improves as more lab data becomes available over time, showing the potential clinical value of a dynamic prediction tool.

Firstly, by modeling multi-class trajectories of blood markers, we provide evidence that there are different temporal pattern trajectories after spine trauma and SCI and that those are, for the most part, non-linear and non-Gaussian responses. Our work distinguishes itself by tackling the multidimensionality, temporality, asynchrony, non-linearity, and heterogeneity inherent in routine blood data—a significant leap from previous studies that only touched on these issues in isolation, which may affect the utility of the models.^37,38^ Considering these factors into the modeling requires a big sample size, which can be challenging in SCI research. Nonetheless, we demonstrated that model development using EHR is feasible and that these models can be generalized to external cohorts with utility for predicting tasks. This should open new exciting venues for SCI research and other medical context since the knowledge gained from EHR can be transferable.

Secondly, using the posterior probability of the modeled class trajectories, we trained classification models with decent performance to detect in-hospital mortality as early as one day after hospital admission. Prediction performance increased as more temporal data was considered for predicting trajectories. This suggests that estimating the probability of which trajectory of blood marker changes patients will follow can serve as predictors in dynamic prognostic neurological outcome models or patient mortality.

Additionally, we fit models to predict whether a patient has an SCI after spine trauma, which could be useful for comatose or obtunded patients when voluntary motor or sensory function cannot be assessed. Prediction performance was better in the case of predicting in-hospital mortality than presence of SCI likely because spine trauma is a significant traumatic event, usually associated with polytraumatic processes (e.g., non-spine fractures) that trigger multiple systemic pathophysiological changes. The addition of other predictors such as injury characteristics could improve classification performance.

We demonstrated that early prediction of marker trajectory in an external cohort (TRACK-SCI patients) has prediction utility. This signifies that dynamic models based on real-world data can be generalized to data collected in other contexts, such as different hospitals. We also found that predicting neurological deficits using only laboratory data is possible, and the prediction improves as more data is included over time in determining patient trajectory. These results mark the first step toward developing a lab-based, dynamic prediction tool for SCI. Future studies will focus on refining key inputs and methods toward the development and implementation of clinical-decision support tools for dynamic prediction.

### Limitations

There are a few limitations to the present work. We limited the analysis to the first 21 days post-injury due to a rapid decrease in sample size beyond this period. However, some patients stayed longer in the hospital. Since the length of hospital stay in SCI is related to patient severity^39^, other trajectories may emerge when more extended periods are considered.

Additionally, our focus was on the most common routine blood markers, potentially excluding other predictors like differential blood white cell counts, which have shown distinct associations with SCI.^17^ These markers were not included due to sample size constraints. Furthermore, since we focused on routine blood markers available in real-world data, we did not include prominent protein biomarkers in neurotrauma, such as the Glial Fibrillary Acidic Protein (GFAP).^40–43^ However, our methodology is adaptable for future inclusion of these and other markers, possibly enhancing predictive accuracy.

Lastly, we focused on the prediction task, hypothesizing that the derived trajectories reflect underlying pathophysiological events. A deeper analysis of the clinical features of each trajectory subpopulation could improve understanding of patient phenotypes and guide the development of better prediction models by identifying early predictors of progressive neurological deficit due to SCI as well as additional stratifying factors.

## Conclusion

We demonstrated the utility of modeling heterogeneous temporal trends of blood markers collected in real-world scenarios to predict diverse diagnostic and prognostic tasks in spine trauma and SCI patients. Longitudinal finite mixture models are powerful tools to describe non-linear multi-class trajectories of blood markers, potentially capturing latent pathophysiological events. Given that SCI and other neurological pathologies evolve, studying the non-linear dynamic changes of any biomarker level is more valuable than considering predictions with a static cut-off level.

The dynamic prediction is promising. By modeling marker trajectories, we can predict a patient’s future pathophysiological changes as early as one day after hospital arrival. This allows for predictive models in diagnostic and prognostic decision-making. These results suggest that real-time prediction models using in-hospital data could support planning and execution of SCI patient management. Future work should focus on implementing these models.

## Data Availability

MIMIC-III and MIMIC-IV are available online under data use agreement through the PhysioNet project. TRACK-SCI are available through a collaboration agreement.

## Acknowledgements

This work has been funded by Wings for Life Foundation to A. Torres-Espin, U.S. Department of Defense (W81XWH-13-1-0297 and W81XWH-16-1-0497), and Craig H. Neilsen Foundation (University of California, San Francisco, Spinal Cord Injury Center of Excellence special project award) to M.S. Beattie. D. Fernández is a Serra-Húnter Fellow, a member of the Centro de Investigación Biomédica en Red de Salud Mental (Instituto de Salud Carlos III), and his work has been supported by the Ministerio de Ciencia e Innovación y Universidades (Spain) [PID2023-148033OB-C21], and by grant 2021 SGR 01421 (GRBIO) administrated by the Departament de Recerca i Universitats de la Generalitat de Catalunya (Spain).

## References

1. James SL, Theadom A, Ellenbogen RG, et al. Global, regional, and national burden of traumatic brain injury and spinal cord injury, 1990–2016: a systematic analysis for the Global Burden of Disease Study 2016. The Lancet Neurology. 2019;18(1):56–87. doi:10.1016/S1474-4422(18)30415-0

2. Merritt CH, Taylor MA, Yelton CJ, Ray SK. Economic impact of traumatic spinal cord injuries in the United States. Neuroimmunology and neuroinflammation. 2019;6.

3. Failli V, Kopp MA, Gericke C, et al. Functional neurological recovery after spinal cord injury is impaired in patients with infections. Brain: A Journal of Neurology. 2012;135(Pt 11):3238–3250. doi:10.1093/brain/aws267

4. Fouad K, Popovich PG, Kopp MA, Schwab JM. The neuroanatomical-functional paradox in spinal cord injury. Nature Reviews Neurology. 2021;17(1):53–62. doi:10.1038/s41582-020-00436-x

5. Jogia T, Kopp MA, Schwab JM, Ruitenberg MJ. Peripheral white blood cell responses as emerging biomarkers for patient stratification and prognosis in acute spinal cord injury. Current Opinion in Neurology. 2021;34(6):796–803. doi:10.1097/WCO.0000000000000995

6. Liebscher T, Ludwig J, Lübstorf T, et al. Cervical Spine Injuries with Acute Traumatic Spinal Cord Injury: Spinal Surgery Adverse Events and Their Association with Neurological and Functional Outcome. Spine. 2022;47(1):E16–E26. doi:10.1097/BRS.0000000000004124

7. Khorasanizadeh M, Yousefifard M, Eskian M, et al. Neurological recovery following traumatic spinal cord injury: a systematic review and meta-analysis. Journal of Neurosurgery Spine. Published online February 2019:1–17. doi:10.3171/2018.10.SPINE18802

8. Albayar AA, Roche A, Swiatkowski P, et al. Biomarkers in spinal cord injury: prognostic insights and future potentials. Frontiers in Neurology. 2019;10:27.

9. Ruiz IA, Squair JW, Phillips AA, et al. Incidence and natural progression of neurogenic shock after traumatic spinal cord injury. Journal of Neurotrauma. 2018;35(3):461–466.

10. Haefeli J, Mabray MC, Whetstone WD, et al. Multivariate analysis of MRI biomarkers for predicting neurologic impairment in cervical spinal cord injury. American Journal of Neuroradiology. 2017;38(3):648–655.

11. Talbott JF, Whetstone WD, Readdy WJ, et al. The Brain and Spinal Injury Center score: a novel, simple, and reproducible method for assessing the severity of acute cervical spinal cord injury with axial T2-weighted MRI findings. Journal of Neurosurgery Spine. 2015;23(4):495–504. doi:10.3171/2015.1.SPINE141033

12. Hawryluk G, Whetstone W, Saigal R, et al. Mean arterial blood pressure correlates with neurological recovery after human spinal cord injury: analysis of high frequency physiologic data. Journal of neurotrauma. 2015;32(24):1958–1967.

13. Squair JW, Bélanger LM, Tsang A, et al. Spinal cord perfusion pressure predicts neurologic recovery in acute spinal cord injury. Neurology. 2017;89(16):1660–1667.

14. Torres-Espín A, Haefeli J, Ehsanian R, et al. Topological network analysis of patient similarity for precision management of acute blood pressure in spinal cord injury. Elife. 2021;10:e68015.

15. Brown SJ, Harrington GMB, Hulme CH, et al. A Preliminary Cohort Study Assessing Routine Blood Analyte Levels and Neurological Outcome after Spinal Cord Injury. Journal of Neurotrauma. 2020;37(3):466–480. doi:10.1089/neu.2019.6495

16. Harrington GMB, Cool P, Hulme C, et al. Routinely Measured Hematological Markers Can Help to Predict American Spinal Injury Association Impairment Scale Scores after Spinal Cord Injury. Journal of Neurotrauma. 2021;38(3):301–308. doi:10.1089/neu.2020.7144

17. Jogia T, Lübstorf T, Jacobson E, et al. Prognostic value of early leukocyte fluctuations for recovery from traumatic spinal cord injury. Clinical and Translational Medicine. 2021;11(1):e272. doi:10.1002/ctm2.272

18. Kwon BK, Stammers AMT, Belanger LM, et al. Cerebrospinal fluid inflammatory cytokines and biomarkers of injury severity in acute human spinal cord injury. Journal of Neurotrauma. 2010;27(4):669–682. doi:10.1089/neu.2009.1080

19. Kwon BK, Bloom O, Wanner IB, et al. Neurochemical biomarkers in spinal cord injury. Spinal Cord. 2019;57(10):819–831. doi:10.1038/s41393-019-0319-8

20. Kyritsis N, Torres-Espín A, Schupp PG, et al. Diagnostic blood RNA profiles for human acute spinal cord injury. Journal of Experimental Medicine. 2021;218(3):e20201795.

21. Leister I, Linde LD, Vo AK, et al. Routine Blood Chemistry Predicts Functional Recovery After Traumatic Spinal Cord Injury: A Post Hoc Analysis. Neurorehabilitation and Neural Repair. 2021;35(4):321–333. doi:10.1177/1545968321992328

22. Bourguignon L, Vo AK, Tong B, et al. Natural Progression of Routine Laboratory Markers after Spinal Trauma: A Longitudinal, Multi-Cohort Study. Journal of Neurotrauma. 2021;38(15):2151–2161. doi:10.1089/neu.2021.0012

23. Furlan JC, Krassioukov AV, Fehlings MG. Hematologic Abnormalities Within the First Week After Acute Isolated Traumatic Cervical Spinal Cord Injury: A Case-Control Cohort Study. Spine. 2006;31(23):2674–2683. doi:10.1097/01.brs.0000244569.91204.01

24. Johnson AE, Pollard TJ, Shen L, et al. MIMIC-III, a freely accessible critical care database. Scientific data. 2016;3(1):1–9.

25. Tsolinas RE, Burke JF, DiGiorgio AM, et al. Transforming Research and Clinical Knowledge in Spinal Cord Injury (TRACK-SCI): an overview of initial enrollment and demographics. Neurosurgical Focus. 2020;48(5):E6.

26. Goldberger AL, Amaral LA, Glass L, et al. PhysioBank, PhysioToolkit, and PhysioNet: components of a new research resource for complex physiologic signals. circulation. 2000;101(23):e215–e220.

27. World Health Organization. International Statistical Classification of Diseases and Related Health Problems: Alphabetical Index. Vol 3. World Health Organization; 2004.

28. Roberts TT, Leonard GR, Cepela DJ. Classifications In Brief: American Spinal Injury Association (ASIA) Impairment Scale. Clinical Orthopaedics and Related Research. 2017;475(5):1499–1504. doi:10.1007/s11999-016-5133-4

29. van der Nest G, Lima Passos V, Candel MJJM, van Breukelen GJP. An overview of mixture modelling for latent evolutions in longitudinal data: Modelling approaches, fit statistics and software. Advances in Life Course Research. 2020;43:100323. doi:10.1016/j.alcr.2019.100323

30. Proust-Lima C, Philipps V, Liquet B. Estimation of Extended Mixed Models Using Latent Classes and Latent Processes: The R Package lcmm. Journal of Statistical Software. 2017;78:1–56. doi:10.18637/jss.v078.i02

31. Schwarz G. Estimating the Dimension of a Model. The Annals of Statistics. 1978;6(2):461–464. doi:10.1214/aos/1176344136

32. Nagin D. Group-Based Modeling of Development. Harvard University Press; 2005.

33. Friedman J, Hastie T, Tibshirani R. Regularization paths for generalized linear models via coordinate descent. Journal of statistical software. 2010;33(1):1.

34. Kuhn M. Caret: classification and regression training. Astrophysics Source Code Library. Published online 2015:ascl-1505.

35. Le Gall JR, Lemeshow S, Saulnier F. A new simplified acute physiology score (SAPS II) based on a European/North American multicenter study. Jama. 1993;270(24):2957–2963.

36. Pickles A, Croudace T. Latent mixture models for multivariate and longitudinal outcomes. Statistical Methods in Medical Research. 2010;19(3):271–289. doi:10.1177/0962280209105016

37. Jolliffe IT, Cadima J. Principal component analysis: a review and recent developments. Philosophical transactions of the royal society A: Mathematical, Physical and Engineering Sciences. 2016;374(2065):20150202.

38. Huie JR, Diaz-Arrastia R, Yue JK, et al. Testing a multivariate proteomic panel for traumatic brain injury biomarker discovery: a TRACK-TBI pilot study. Journal of neurotrauma. 2019;36(1):100–110.

39. Jang HJ, Park J, Shin HI. Length of hospital stay in patients with spinal cord injury. Annals of Rehabilitation Medicine. 2011;35(6):798.

40. Korley FK, Jain S, Sun X, et al. Prognostic value of day-of-injury plasma GFAP and UCH-L1 concentrations for predicting functional recovery after traumatic brain injury in patients from the US TRACK-TBI cohort: an observational cohort study. The Lancet Neurology. 2022;21(9):803–813. doi:10.1016/S1474-4422(22)00256-3

41. Abdelhak A, Foschi M, Abu-Rumeileh S, et al. Blood GFAP as an emerging biomarker in brain and spinal cord disorders. Nat Rev Neurol. 2022;18(3):158–172. doi:10.1038/s41582-021-00616-3

42. Okonkwo DO, Yue JK, Puccio AM, et al. GFAP-BDP as an Acute Diagnostic Marker in Traumatic Brain Injury: Results from the Prospective Transforming Research and Clinical Knowledge in Traumatic Brain Injury Study. Journal of Neurotrauma. 2013;30(17):1490. doi:10.1089/neu.2013.2883

43. Stukas S, Cooper J, Gill J, et al. Association of CSF and Serum Neurofilament Light and Glial Fibrillary Acidic Protein, Injury Severity, and Outcome in Spinal Cord Injury. Neurology. 2023;100(12):e1221. doi:10.1212/WNL.0000000000206744

