## Supplementary Material for "Modeling trajectories of routine blood tests as dynamic biomarkers for outcome in spinal cord injury"

**eMethods.**

**eTable 1**. Cross-tabulation of laboratory analytes per category and fluid sample

**eTable 2.** Final selected GMM models

**eTable 3.** Univariate comparisons between clusters of Anion Gap and Chloride.

**eTable 4**. Univariate comparisons between clusters of Glucose and Magnesium.

**eTable 5.** Univariate comparisons between clusters of Phosphate and Potassium.

**eTable 6.** Univariate comparisons between clusters of Sodium and Urea Nitrogen
**eTable 7.** Univariate comparisons between clusters of Hematocrit and Hemoglobin.
**eTable 8.** Univariate comparisons between clusters of MCH and MCHC.

**eTable 9.** Univariate comparisons between clusters of MCV and Platelet Count.

**eTable 10.** Univariate comparisons between clusters of RDW and Red Blood Cells.

**eTable 11.** Univariate comparisons between clusters of White blood cells.

**eFigure 1**. Spaghetti plots for the raw data of the modeling set of analytes (20 most common).

**eFigure 2**. Spaghetti plots for the outlier-cleaned modeling set of laboratory analytes for the first 21 days after admission.

**eFigure 3.** Spaghetti plots for the outlier-cleaned TRACK-SCI minimal set of laboratory analytes for the first 21 days after admission.

**eFigure 4**. Length of stay distribution.

**eFigure 5**. Example of model fit plots for two different types of model selection "patterns".

**eFigure 6.** Average posterior probability of assignment (APPA) for each trajectory class and marker at each time point window.

**eFigure 7.** Average posterior probability of assignment (APPA) for each trajectory class and marker at each time point window in TRACJ_SCI.

**eFigure 8**. PR-AUC performance of dynamic predictions on the train data.

**eFigure 9**. ROC-AUC performance of dynamic predictions on the train data.

**eFigure 10**. ROC-AUC performance of dynamic predictions on the out-train data.

**eFigure 11**. ROC-AUC performance of dynamic predictions on the train data for ICU patients.

**eFigure 12**. PR-AUC performance of dynamic predictions on the train data for ICU patients.

**eFigure 13**. ROC-AUC performance of dynamic predictions on the out-train data for ICU patients.

**eFigure 14**. PR-AUC performance of dynamic predictions on the out-train data for ICU patients.

**eFigure 15**. Variable importance for models with posterior probability of trajectory classification only.

**eFigure 16**. Variable importance for models with posterior probability of trajectory classification and summary statistics of biomarkers.

**eFigure 17**. Variable importance for models with posterior probability of trajectory classification, summary statistics and baseline predictors.

### eMethods.

#### Cohort selection based on ICD9/ICD10

International Classification of Diseases (ICD9/ICD10) diagnostic codes are for emergency hospital admissions. These include all codes of the series ICD9: 951, 953, 806 and 805, and ICD10: S110, S111, S112, S113, S114, S115, S116, S118, S119, S140, S141, S142, S210, S240, S241, S310, S311, S340, S341, S342, S343.

#### Demographics and hospital stay characteristics.

For MIMIC-III, patient age was derived by subtracting the admission date (ADMITTIME) from the birth date (DOB). In MIMIC, the actual date of birth is not accessible; instead, a shifted version is provided. This date shift is consistent across all timestamps for each patient, allowing for the calculation of age by subtracting the shifted date of birth from the time of admission. Gender did not require any data cleaning. To align the two databases, ethnicity, insurance, admission type, and locations were standardized by merging similar categories to avoid information loss. The analysis was restricted to emergency admissions. The length of stay was determined by the difference between admission time and discharge time (DISCHTIME).

#### Exploratory data analysis and modeling data

We excluded laboratory values with missing Logical Observation Identifier Names and Codes. A total of 413 distinct laboratory markers were observed in the data. The number of markers per category and fluid sample are cross tabulated in eTable 1. Of the 413, 157 were categorized as hematology, 161 as chemistry, and 25 as blood gases. Regarding the fluid sample, the fluid with more distinct markers was blood, with blood chemistry and hematology being the two categories with more markers. The 20 most common markers, found in 90-98% of patients, were all from hematology and chemistry, measured in blood. Less than 70% of patients had the remaining markers, with 341 markers present in 10% or fewer patients.

Temporal spaghetti and marginal density plots were generated to understand the amount of available data for the laboratory markers, the underlying distribution, the potential presence of outliers, and non-linearities. Spaghetti plots of the marker modeling set over time from the date of hospital arrival for MIMIC patients are shown in eFigure 1. As expected by the nature of the data, time is asynchronous, meaning that markers were obtained for each patient at different timepoints in non-regular intervals. No apparent trends were observable from the plots. We can observe fluctuations over time, with, in general, a high dynamic range early after admission that reduces as time progresses. The number of subjects with data in each marker also reduces over time, with very few subjects with data beyond 50 days (eFigure 2). This can also be confirmed by the distribution of length of stay, where the median time is 4.6 days (first quartile: 2.09 and third quartile: 9.07), and 96.4% of the cohort was discharged before 28 days in the hospital.

We can also observe subtle spikes in the data from the spaghetti plots, probably caused by errors in the original MIMIC dataset. These spikes create extreme values in the marginal distributions for each marker in the modeling set (eFigure 2). Using the measurement range values, we can confirm that some of these values are outliers. For example, values of 0 in MCH, as we see in the plot, are not possible (as MCH is the ratio of hemoglobin to red blood cells). Although the number of spikes is small, we performed a data filter to detect and discard those highly likely observations of outliers. First, 0 values were excluded as these are unprovable in any of the modeling set of markers. Next, we applied a variation of John Tukey's rule for outlier determination as proportional to the interquartile (IQR) range. [1] In this case, we filtered out extreme values with $Lower\ limit = quantile_{10} - 1.5IQR$ and $Upper\ limit = quantile_{80} + 1.5IQR$ on the subject-specific marginal distribution for each marker. Note that 10% and 80% quantiles instead of Tukey's 15% and 75% were used to be more permissive, especially for skewed distributions where Tukey's can be too restrictive. [2] This results in less extreme values per marker. Finally, given the high drop in the number of patients with data available beyond the first few weeks, we limited the set of markers for modeling to 21 days from hospital admission (eFigure 3). The same pre-processing conducted for MIMIC was done for the TRACK-SCI lab values. The spaghetti plots for the outlier-cleaned TRACK-SCI minimal set of laboratory analytes for the first 21 days after admission are shown in eFigure 4.

#### Multi-class trajectory modeling

Researchers have previously recommended a staged workflow because of the computational time it takes to run these models and the difficulty of prioritizing model selection. It starts by running a search of the parameter space using LCGA, discarding the models that are not a good fit, and adjusting other parameters before incorporating random effects. We followed such a strategy, performing a grid search for each one of the modeling markers for the number of classes (1 to 5), the degree of polynomials (1 to 3), and three distinct link functions: linear or identity for Gaussian continuous responses, and beta and I-spline to model non-Gaussian continuous responses. A total of 900 models were specified. For model selection, we used BIC, ICL, and APPA, prioritizing lower BIC and ICL values and an APPA above 0.7. Each class was required to represent at least 1% of subjects.

Plots of the model criteria for these models for red blood cell count and potassium are provided in eFigure 5. An interpretation of the model fit selection would go as follows. For red blood cell count, the polynomial degree is the dominant determinant of better model fit by both BIC and ICL. In these cases, we conclude that non-linear trends are a better fit and that the type of link function is almost irrelevant. This informs that assuming a Gaussian response is acceptable for markers with this pattern of model fit. Regarding the number of classes, for red blood cell count, BIC reduces progressively with the increase of classes, while ICL presents a more prominent drop at two classes but with a more stable value afterward. APPA has a more linear reduction as the number of classes increases. Given that APPA reached acceptable levels in most cases, we used BIC and ICL as the main decision-making tools for this initial step, leaving APPA as a secondary factor. Therefore, we concluded that a linear link and a polynomial of order 3 are best for this variable, selecting between 2 to 4 trajectory classes for modeling red blood cell count. In a different example, we can look at Potassium, where both the polynomial order and the shape of the link function are important for model fit. In this case, we selected models with polynomials of order 3, link function of type beta or spline, and 2 or 3 trajectory classes for further exploration.

In summary, most markers show that at least 2, with a median of 3, trajectory classes are needed to model the data, indicative of heterogeneity in the selected cohort. In addition, most models presented a better fit with a polynomial transformation of degree 2 or 3, indicative that non-linear trends are present in the data. Finally, with the exception of bicarbonate, chloride, hematocrit, hemoglobin, MCHC, and red blood cell count, all other marker data were better fitted to models with non-linear link functions (beta or I-spline), which suggests that blood markers are better modeled by non-Gaussian conditional distributions.

After model exploration, we selected those models that showed a better fit based on BIC, ICL, and APPA (eTable 2). There was a clear best choice for some markers, while the selection was more subjective for others. In those cases, we selected different models for the next step. We then went through a similar process, using GMMs, but now with the inclusion of the random effects. In addition, instead of polynomial transformations for the time variable, to relax some of the polynomial geometrical constrictions, we used a natural spline to model non-linear trends, with degrees determined through the exact search as before. A total of 122 models were specified. The posterior probability of class assignment (PPA) for each marker and subject was calculated, and class membership was assigned using maximum a posteriori probability estimates. Those posterior probabilities were used as predictors in an Elastic Net model for classification. PPA for each marker were high as early as 1 day of hospital arrival, with an average of 0.93 (SD: 0.12) across markers, classes, and timepoints (eFigures 6-7).

Univariate analysis comparing class trajectories for each marker for different demographics and clinical characteristics can be found in the eTables 3 to 11. In brief, significant differences were observed in age, the proportion of patients dead during hospitalization, and the number of diagnostics for most markers. The class trajectories with lower subject numbers in anion gap, chloride, glucose, magnesium, phosphate, potassium, sodium, and blood urea nitrogen were associated with higher in-hospital mortality rates and a higher number of diagnostics. Except for sodium, those smaller class trajectories were generally characterized by higher values of the markers early after hospital arrival, with major non-linear dynamic changes over the first three weeks compared to the class with the most patients (Fig. 2f). Of the chemical markers, only glucose and sodium showed differences in the proportion of males and females between trajectory classes. In the case of hematology markers, class 1 for hematocrit, hemoglobin, and red blood cell counts showed higher initial marker values with a posterior drop was associated with a lower mortality rate and the number of diagnostics than classes 2 and 3. Length of stay differences can be found between classes of glucose, hematocrit, hemoglobin, platelet count, and red blood cells. Finally, the proportion of patients in each cohort group (SCI Fracture, SCI noFracture, and Spine Trauma) varied across classes for glucose, hematocrit, hemoglobin, platelet count, red blood cell counts, and white blood cell counts.

**Machin Learning Classifiers and Model Performance**

The Simplified Acute Physiology Score (SAPS), originally developed in 1984, combines 13 physiological measures and the patient's age to assess the mortality risk for ICU patients. By 1993, the model was updated to SAPS II, incorporating 17 factors in total: 12 physiological metrics, age, admission type (whether it was a scheduled surgery, an emergency surgery, or a medical reason), and three variables for pre-existing conditions (AIDS, metastatic cancer, and blood cancers). For the physiological variables, the worst value during the first 24 hours of ICU admission is used for the calculation.[3] SAPS II was further modified in 2005, and six more criteria for assessment upon admission, including age, gender, the duration of hospital stay before ICU admission, the patient's location prior to ICU, the clinical reason for admission, and whether there was a drug overdose involved was added. The latest version is used in this article. We compute scores using SQL code publicly available on GitHub. (https://github.com/MIT-LCP/mimic-code/blob/main/mimic-iii/concepts/severityscores/sapsii.sql)

When comparing model performance, note that SAPS II incorporates baseline data, physiological metrics, and their summary statistics. Therefore, its performance should be evaluated against models that also integrate trajectories, summary statistics, and baseline information.

**Variable Importance**

We assessed variable importance based on the magnitude of these standardized coefficients, normalizing the scores so the most significant variable receives a score of 100, relative to others. Variables with mean importance of more than 40 we demonstrated for each Experiment and cut-off in eFigures 13-15.

### eTables

#### eTable 1. Cross-tabulation of laboratory analytes per category and fluid

| **Fluid sample \ Category** | Blood Gas | Chemistry | Hematology | **Total** |
|---|---|---|---|---|
| Ascites | 0 | 9 | 13 | **22** |
| Blood | 24 | 104 | 71 | **199** |
| Cerebrospinal Fluid (CSF) | 0 | 3 | 13 | **16** |
| Joint Fluid | 0 | 1 | 11 | **12** |
| Other Body Fluid | 1 | 9 | 14 | **24** |
| Pleural | 0 | 8 | 15 | **23** |
| Urine | 0 | 27 | 20 | **47** |
| *Unknown* | 0 | 0 | 0 | **70** |
| **Total** | **25** | **161** | **157** | **413** |

#### eTable 2. Final selected GMM models

| **Analyte** | **k** | **link** | **np** | **d** | **BIC** | **ICL** | **APP** | **%clas** | **%clas** | **%clas** | **%clas** | **%clas** |
|---|---|---|---|---|---|---|---|---|---|---|---|---|
| Anion Gap | 2 | beta | 22 | 3 | 104781. | 99681.8 | 0.98 | 2.00 | 98.00 | NA | NA | NA |
| Bicarbonate | 1 | linea | 15 | 3 | 111559.0 | 111559. | 1.00 | 100.00 | NA | NA | NA | NA |
| Calcium, Total | 1 | linea | 10 | 2 | 24868.3 | 24868.3 | 1.00 | 100.00 | NA | NA | NA | NA |
| Chloride | 2 | beta | 22 | 3 | 127905. | 122725. | 1.00 | 0.85 | 99.15 | NA | NA | NA |
| Glucose | 2 | beta | 11 | 1 | 222867. | 218145. | 0.91 | 10.46 | 89.54 | NA | NA | NA |
| Hematocrit | 3 | linea | 25 | 3 | 138572. | 134284. | 0.82 | 31.93 | 13.51 | 54.56 | NA | NA |
| Hemoglobin | 3 | linea | 25 | 3 | 64672.1 | 60404.3 | 0.82 | 26.74 | 11.37 | 61.89 | NA | NA |
| Magnesium | 2 | splin | 23 | 3 | -4861.69 | - | 0.97 | 1.95 | 98.05 | NA | NA | NA |
| MCH | 3 | linea | 18 | 2 | 46805.7 | 41775.4 | 0.97 | 94.97 | 1.84 | 3.19 | NA | NA |
| MCHC | 2 | linea | 14 | 2 | 59109.6 | 54145.6 | 0.95 | 96.54 | 3.46 | NA | NA | NA |
| MCV | 2 | beta | 16 | 2 | 97687.9 | 92518.5 | 0.99 | 98.92 | 1.08 | NA | NA | NA |
| Phosphate | 2 | beta | 16 | 2 | 50457.5 | 45400.0 | 0.99 | 2.74 | 97.26 | NA | NA | NA |
| Platelet Count | 2 | beta | 22 | 3 | 248904. | 246232. | 0.51 | 35.65 | 64.35 | NA | NA | NA |
| Potassium | 2 | beta | 22 | 3 | 23972.6 | 18936.7 | 0.97 | 3.11 | 96.89 | NA | NA | NA |
| RDW | 3 | beta | 20 | 2 | 34238.7 | 29400.8 | 0.93 | 8.99 | 87.09 | 3.92 | NA | NA |
| Red Blood | 3 | linea | 25 | 3 | 13818.2 | 9586.07 | 0.81 | 18.94 | 10.95 | 70.11 | NA | NA |
| Sodium | 2 | beta | 16 | 2 | 129149. | 124014. | 0.99 | 97.35 | 2.65 | NA | NA | NA |
| Urea Nitrogen | 2 | beta | 16 | 2 | 146168. | 141216. | 0.95 | 7.65 | 92.35 | NA | NA | NA |
| White Blood | 3 | beta | 27 | 3 | 109955. | 104834. | 0.98 | 0.85 | 98.08 | 1.08 | NA | NA |

### eTable 3. Univariate comparisons between clusters of Anion Gap and Chloride

| Characteristic | Anion Gap 1 N = 52 | Anion Gap 2 N = 2548 | Anion Gap p-value | Anion Gap q-value | Chloride 1 N = 22 | Chloride 2 N = 2578 | Chloride p-value | Chloride q-value |
|---|---|---|---|---|---|---|---|---|
| Age | 76 (58, 83) | 62 (43, 79) | <0.001 | **<0.001** | 28 (22, 55) | 63 (43, 80) | <0.001 | **<0.001** |
| Gender | | | 0.6 | 0.7 | | | 0.3 | 0.4 |
| F | 19 (37%) | 1,036 (41%) | | | 6 (27%) | 1,049 (41%) | | |
| M | 33 (63%) | 1,512 (59%) | | | 16 (73%) | 1,529 (59%) | | |
| Ethnicity | | | 0.7 | 0.7 | | | 0.4 | 0.4 |
| ASIAN | 2 (4.7%) | 47 (2.1%) | | | 0 (0%) | 49 (2.2%) | | |
| BLACK | 1 (2.3%) | 124 (5.6%) | | | 0 (0%) | 125 (5.6%) | | |
| HISPANIC | 2 (4.7%) | 96 (4.3%) | | | 1 (6.7%) | 97 (4.3%) | | |
| MULTI RACE | 0 (0%) | 6 (0.3%) | | | 0 (0%) | 6 (0.3%) | | |
| OTHER | 1 (2.3%) | 103 (4.7%) | | | 2 (13%) | 102 (4.6%) | | |
| WHITE | 37 (86%) | 1,833 (83%) | | | 12 (80%) | 1,858 (83%) | | |
| Unknown | 9 | 339 | | | 7 | 341 | | |
| Cohort | | | 0.051 | 0.090 | | | 0.7 | 0.7 |
| SCI Fracture | 2 (3.8%) | 380 (15%) | | | 4 (18%) | 378 (15%) | | |
| SCI noFracture | 3 (5.8%) | 121 (4.7%) | | | 1 (4.5%) | 123 (4.8%) | | |
| Spine Trauma | 47 (90%) | 2,047 (80%) | | | 17 (77%) | 2,077 (81%) | | |
| Length of stay (days) | 7 (5, 11) | 7 (4, 13) | 0.6 | 0.7 | 9 (4, 14) | 7 (5, 12) | 0.2 | 0.4 |
| Unknown | 0 | 1 | | | 0 | 1 | | |
| Died in hospital | 20 (38%) | 169 (6.6%) | <0.001 | **<0.001** | 14 (64%) | 175 (6.8%) | <0.001 | **<0.001** |
| Number of diagnostics | 26 (17, 30) | 13 (9, 19) | <0.001 | **<0.001** | 14 (9, 26) | 13 (9, 20) | 0.13 | 0.3 |

### eTable 4. Univariate comparisons between clusters of Glucose and Magnesium

| | Glucose | | | | Magnesium | | | |
|---|---|---|---|---|---|---|---|---|
| **Characteristic** | **1**<br>**N = 272** | **2**<br>**N = 2328** | **p-value** | **q-value** | **1**<br>**N = 50** | **2**<br>**N = 2513** | **p-value** | **q-value** |
| Age | 69 (56, 80) | 62 (41, 80) | <0.001 | **<0.001** | 62 (49, 78) | 62 (43, 79) | 0.7 | >0.9 |
| Gender | | | 0.013 | **0.015** | | | >0.9 | >0.9 |
| F | 91 (33%) | 964 (41%) | | | 20 (40%) | 1,017 (40%) | | |
| M | 181 (67%) | 1,364 (59%) | | | 30 (60%) | 1,496 (60%) | | |
| Ethnicity | | | 0.3 | 0.3 | | | 0.5 | 0.9 |
| ASIAN | 8 (3.5%) | 41 (2.0%) | | | 1 (2.7%) | 48 (2.2%) | | |
| BLACK | 12 (5.2%) | 113 (5.6%) | | | 3 (8.1%) | 121 (5.6%) | | |
| HISPANIC | 14 (6.1%) | 84 (4.2%) | | | 3 (8.1%) | 94 (4.3%) | | |
| MULTI RACE | 0 (0%) | 6 (0.3%) | | | 0 (0%) | 6 (0.3%) | | |
| OTHER | 14 (6.1%) | 90 (4.4%) | | | 2 (5.4%) | 99 (4.5%) | | |
| WHITE | 181 (79%) | 1,689 (83%) | | | 28 (76%) | 1,811 (83%) | | |
| Unknown | 43 | 305 | | | 13 | 334 | | |
| Cohort | | | 0.002 | **0.003** | | | 0.2 | 0.6 |
| SCI Fracture | 50 (18%) | 332 (14%) | | | 10 (20%) | 371 (15%) | | |
| SCI noFracture | 23 (8.5%) | 101 (4.3%) | | | 4 (8.0%) | 119 (4.7%) | | |
| Spine Trauma | 199 (73%) | 1,895 (81%) | | | 36 (72%) | 2,023 (81%) | | |
| Length of stay (days) | 9 (5, 18) | 7 (4, 12) | <0.001 | **<0.001** | 8 (5, 14) | 7 (5, 13) | >0.9 | >0.9 |
| Unknown | 0 | 1 | | | 0 | 1 | | |
| Died in hospital | 38 (14%) | 151 (6.5%) | <0.001 | **<0.001** | 13 (26%) | 176 (7.0%) | <0.001 | **<0.001** |
| Number of diagnostics | 17 (11, 26) | 13 (9, 19) | <0.001 | **<0.001** | 20 (10, 27) | 13 (9, 19) | <0.001 | **<0.001** |

### eTable 5. Univariate comparisons between clusters of Phosphate and Potassium

| Characteristic | Phosphate<br>1<br>N = 72 | <br>2<br>N = 2481 | <br>p-value | <br>q-value | Potassium<br>1<br>N = 81 | <br>2<br>N = 2521 | <br>p-value | <br>q-value |
|---|---|---|---|---|---|---|---|---|
| Age | 70 (56, 81) | 62 (43, 79) | 0.006 | **0.013** | 73 (60, | 62 (43, 79) | <0.001 | **<0.001** |
| Gender | | | >0.9 | >0.9 | | | 0.7 | 0.8 |
| F | 30 (42%) | 1,003 | | | 31 (38%) | 1,024 | | |
| M | 42 (58%) | 1,478 | | | 50 (62%) | 1,497 | | |
| Ethnicity | | | >0.9 | >0.9 | | | 0.3 | 0.5 |
| ASIAN | 2 (3.5%) | 47 (2.2%) | | | 2 (2.8%) | 47 (2.2%) | | |
| BLACK | 2 (3.5%) | 122 (5.7%) | | | 1 (1.4%) | 124 (5.7%) | | |
| HISPANIC | 2 (3.5%) | 94 (4.4%) | | | 1 (1.4%) | 97 (4.4%) | | |
| MULTI | 0 (0%) | 6 (0.3%) | | | 0 (0%) | 6 (0.3%) | | |
| OTHER | 2 (3.5%) | 98 (4.6%) | | | 5 (7.0%) | 99 (4.5%) | | |
| WHITE | 49 (86%) | 1,782 | | | 62 (87%) | 1,810 | | |
| Unknown | 15 | 332 | | | 10 | 338 | | |
| Cohort | | | 0.5 | 0.7 | | | 0.06 | 0.11 |
| SCI Fracture | 7 (9.7%) | 371 (15%) | | | 5 (6.2%) | 377 (15%) | | |
| SCI | 4 (5.6%) | 118 (4.8%) | | | 3 (3.7%) | 122 (4.8%) | | |
| Spine | 61 (85%) | 1,992 | | | 73 (90%) | 2,022 | | |
| Length of stay (days) | 8 (5, 15) | 7 (5, 13) | 0.5 | 0.7 | 8 (5, 13) | 7 (4, 12) | 0.8 | 0.8 |
| Unknown | 0 | 1 | | | 0 | 1 | | |
| Died in hospital | 26 (36%) | 161 (6.5%) | <0.001 | **<0.001** | 15 (19%) | 174 (6.9%) | <0.001 | **0.001** |
| Number of | 24 (17, 33) | 13 (9, 19) | <0.001 | **<0.001** | 21 (14, | 13 (9, 19) | <0.001 | **<0.001** |

**eTable 6. Univariate comparisons between clusters of Sodium and Urea**

| Characteristic | Sodium | | | | Urea Nitrogen | | | |
|---|---|---|---|---|---|---|---|---|
| | **1**<br>**N = 2531** | **2**<br>**N = 69** | **p-value** | **q-value** | **1**<br>**N = 199** | **2**<br>**N = 2401** | **p-value** | **q-value** |
| Age | 62 (43, | 78 (64, 87) | <0.001 | **<0.001** | 78 (69, | 61 (41, 78) | <0.001 | **<0.001** |
| Gender | | | <0.001 | **<0.001** | | | >0.9 | >0.9 |
| F | 1,008 | 47 (68%) | | | 81 (41%) | 974 (41%) | | |
| M | 1,523 | 22 (32%) | | | 118 | 1,427 | | |
| Ethnicity | | | 0.3 | 0.4 | | | 0.058 | 0.1 |
| ASIAN | 46 (2.1%) | 3 (4.5%) | | | 8 (4.4%) | 41 (2.0%) | | |
| BLACK | 122 | 3 (4.5%) | | | 7 (3.9%) | 118 (5.7%) | | |
| HISPANIC | 98 (4.5%) | 0 (0%) | | | 5 (2.8%) | 93 (4.5%) | | |
| MULTI | 6 (0.3%) | 0 (0%) | | | 0 (0%) | 6 (0.3%) | | |
| OTHER | 102 | 2 (3.0%) | | | 3 (1.7%) | 101 (4.9%) | | |
| WHITE | 1,812 | 58 (88%) | | | 157 | 1,713 | | |
| Unknown | 345 | 3 | | | 19 | 329 | | |
| Cohort | | | 0.076 | 0.11 | | | 0.08 | 0.1 |
| SCI | 378 (15%) | 4 (5.8%) | | | 20 (10%) | 362 (15%) | | |
| SCI | 121 | 3 (4.3%) | | | 13 (6.5%) | 111 (4.6%) | | |
| Spine | 2,032 | 62 (90%) | | | 166 | 1,928 | | |
| Length of stay | 7 (5, 13) | 6 (4, 11) | 0.6 | 0.6 | 7 (4, 11) | 7 (5, 13) | 0.082 | 0.1 |
| Unknown | 1 | 0 | | | 0 | 1 | | |
| Died in hospital | 179 | 10 (14%) | 0.03 | 0.053 | 36 (18%) | 153 (6.4%) | <0.001 | **<0.001** |
| Number of | 13 (9, 19) | 17 (13, 27) | <0.001 | **<0.001** | 20 (14, | 13 (9, 19) | <0.001 | **<0.001** |

### eTable 7. Univariate comparisons between clusters of Hematocrit and Hemoglobin

| | **Hematocrit** | | | | | **Hemoglobin** | | | | |
|---|---|---|---|---|---|---|---|---|---|---|
| **Characteristic** | **1 N = 834** | **2 N = 353** | **3 N = 1425** | **p valu e** | **q valu e** | **1 N = 696** | **2 N = 296** | **3 N = 1,611** | **p value** | **q valu e** |
| Age | 64 (45, | 47 (29, 63) | 66 (47, | <0.00 | **<0.00** | 61 (42, | 44 (29, 60) | 66 (47, | <0.00 | **<0.00** |
| Gender | | | | <0.00 | **<0.00** | | | | <0.00 | **<0.00** |
| F | 295 (35%) | 79 (22%) | 686 (48%) | | | 222 | 59 (20%) | 774 (48%) | | |
| M | 539 (65%) | 274 (78%) | 739 (52%) | | | 474 | 237 (80%) | 837 (52%) | | |
| Ethnicity | | | | 0.5 | 0.5 | | | | 0.2 | 0.2 |
| ASIAN | 21 (2.8%) | 5 (1.8%) | 23 (1.9%) | | | 17 (2.7%) | 4 (1.7%) | 28 (2.0%) | | |
| BLACK | 42 (5.5%) | 12 (4.3%) | 71 (5.8%) | | | 32 (5.1%) | 8 (3.4%) | 85 (6.1%) | | |
| HISPANIC | 37 (4.9%) | 17 (6.1%) | 46 (3.7%) | | | 31 (4.9%) | 14 (6.0%) | 53 (3.8%) | | |
| MULTI | 1 (0.1%) | 1 (0.4%) | 4 (0.3%) | | | 1 (0.2%) | 1 (0.4%) | 4 (0.3%) | | |
| OTHER | 31 (4.1%) | 17 (6.1%) | 58 (4.7%) | | | 29 (4.6%) | 18 (7.7%) | 59 (4.2%) | | |
| WHITE | 625 (83%) | 227 (81%) | 1,026 | | | 517 | 189 (81%) | 1,166 | | |
| Unknown | 77 | 74 | 197 | | | 69 | 62 | 216 | | |
| Cohort | | | | <0.00 | **<0.00** | | | | <0.00 | **<0.00** |
| SCI | 93 (11%) | 94 (27%) | 195 (14%) | | | 81 (12%) | 70 (24%) | 231 (14%) | | |
| SCI | 57 (6.8%) | 5 (1.4%) | 63 (4.4%) | | | 50 (7.2%) | 7 (2.4%) | 68 (4.2%) | | |
| Spine | 684 (82%) | 254 (72%) | 1,167 | | | 565 | 219 (74%) | 1,312 | | |
| Length of stay | 5 (4, 9) | 9 (6, 16) | 8 (5, 14) | <0.00 | **<0.00** | 6 (4, 9) | 8 (5, 16) | 8 (5, 14) | <0.00 | **<0.00** |
| Unknown | 0 | 1 | 0 | | | 0 | 0 | 1 | | |
| Died in hospital | 39 (4.7%) | 29 (8.2%) | 121 | 0.004 | **0.005** | 24 (3.4%) | 29 (9.8%) | 136 | <0.00 | **<0.00** |
| Number of | 12 (8, 18) | 13 (9, 20) | 14 (9, 20) | <0.00 | **<0.00** | 11 (8, 16) | 13 (9, 20) | 15 (9, 20) | <0.00 | **<0.00** |

### eTable 8. Univariate comparisons between clusters of MCH and MCHC

| | MCH | | | | | MCHC | | | |
|---|---|---|---|---|---|---|---|---|---|
| **Characteristic** | **1**<br>**N = 2472** | **2**<br>**N = 48** | **3**<br>**N = 83** | **p value** | **q valu** | **1**<br>**N = 2513** | **2**<br>**N = 90** | **p value** | **q value** |
| Age | 62 (43, 79) | 74 (52, 83) | 64 (46, 80) | 0.01 | **0.023** | 62 (43, 79) | 74 (60, 85) | <0.001 | **<0.00** |
| Gender | | | | 0.3 | 0.3 | | | 0.004 | **0.01** |
| F | 1,002 [illegible] | 15 (31%) | 38 (46%) | | | 1,005 (40%) | 50 (56%) | | |
| M | 1,470 [illegible] | 33 (69%) | 45 (54%) | | | 1,508 (60%) | 40 (44%) | | |
| Ethnicity | | | | <0.001 | **0.003** | | | 0.3 | 0.3 |
| ASIAN | 44 (2.1%) | 0 (0%) | 5 (6.8%) | | | 46 (2.1%) | 3 (3.6%) | | |
| BLACK | 112 (5.2%) | 1 (2.4%) | 12 (16%) | | | 116 (5.3%) | 9 (11%) | | |
| HISPANIC | 89 (4.2%) | 1 (2.4%) | 8 (11%) | | | 95 (4.4%) | 3 (3.6%) | | |
| MULTI RACE | 6 (0.3%) | 0 (0%) | 0 (0%) | | | 6 (0.3%) | 0 (0%) | | |
| OTHER | 101 (4.7%) | 1 (2.4%) | 4 (5.4%) | | | 102 (4.7%) | 4 (4.8%) | | |
| WHITE | 1,789 [illegible] | 38 (93%) | 45 (61%) | | | 1,807 (83%) | 65 (77%) | | |
| Unknown | 331 | 7 | 9 | | | 341 | 6 | | |
| Cohort | | | | 0.2 | 0.3 | | | 0.3 | 0.3 |
| SCI Fracture | 364 (15%) | 9 (19%) | 9 (11%) | | | 373 (15%) | 9 (10%) | | |
| SCI | 116 (4.7%) | 1 (2.1%) | 8 (9.6%) | | | 119 (4.7%) | 6 (6.7%) | | |
| Spine Trauma | 1,992 (81%) | 38 (79%) | 66 (80%) | | | 2,021 (80%) | 75 (83%) | | |
| Length of stay (days) | 7 (4, 13) | 7 (5, 11) | 7 (4, 12) | 0.8 | 0.8 | 7 (4, 13) | 7 (5, 11) | 0.2 | 0.3 |
| Unknown | 1 | 0 | 0 | | | 1 | 0 | | |
| Died in hospital | 180 (7.3%) | 7 (15%) | 2 (2.4%) | 0.035 | 0.061 | 180 (7.2%) | 9 (10%) | 0.3 | 0.3 |
| Number of | 13 (9, 19) | 16 (11, 22) | 17 (12, 21) | <0.001 | **0.002** | 13 (9, 19) | 21 (17, 28) | <0.001 | **<0.00** |

### eTable 9. Univariate comparisons between clusters of MCV and Platelet Count

| | **MCV** | | | | **Platelet Count** | | | | |
|---|---|---|---|---|---|---|---|---|---|
| **Characteristic** | **1**<br>**N = 2575** | **2**<br>**N = 28** | **p value** | **q valu** | **1**<br>**N = 51** | **2**<br>**N = 65** | **3**<br>**N = 2487** | **p value** | **q value** |
| Age | 62 (43, 80) | 73 (63, 83) | 0.012 | **0.028** | 77 (60, [illegible] | 50 (36, 67) | 62 (43, 80) | <0.001 | **<0.00**[illegible] |
| Gender | | | 0.8 | 0.8 | | | | <0.001 | **<0.00**[illegible] |
| F | 1,043 (41%) | 12 (43%) | | | 36 (71%) | 24 (37%) | 995 (40%) | | |
| M | 1,532 (59%) | 16 (57%) | | | 15 (29%) | 41 (63%) | 1,492 [illegible] | | |
| Ethnicity | | | 0.3 | 0.5 | | | | 0.6 | 0.6 |
| ASIAN | 48 (2.1%) | 1 (4.8%) | | | 0 (0%) | 1 (1.9%) | 48 (2.2%) | | |
| BLACK | 125 (5.6%) | 0 (0%) | | | 3 (6.1%) | 3 (5.8%) | 119 (5.5%) | | |
| HISPANIC | 98 (4.4%) | 0 (0%) | | | 2 (4.1%) | 3 (5.8%) | 93 (4.3%) | | |
| MULTI RACE | 6 (0.3%) | 0 (0%) | | | 0 (0%) | 0 (0%) | 6 (0.3%) | | |
| OTHER | 104 (4.7%) | 2 (9.5%) | | | 0 (0%) | 5 (9.6%) | 101 (4.7%) | | |
| WHITE | 1,854 (83%) | 18 (86%) | | | 44 (90%) | 40 (77%) | 1,788 (83%) | | |
| Unknown | 340 | 7 | | | 2 | 13 | 332 | | |
| Cohort | | | 0.6 | 0.7 | | | | 0.006 | **0.008** |
| SCI Fracture | 377 (15%) | 5 (18%) | | | 7 (14%) | 16 (25%) | 359 (14%) | | |
| SCI noFracture | 125 (4.9%) | 0 (0%) | | | 7 (14%) | 0 (0%) | 118 (4.7%) | | |
| Spine Trauma | 2,073 (81%) | 23 (82%) | | | 37 (73%) | 49 (75%) | 2,010 [illegible] | | |
| Length of stay (days) | 7 (4, 13) | 7 (5, 9) | 0.082 | 0.14 | 9 (4, 12) | 20 (13, 27) | 7 (4, 12) | <0.001 | **<0.00**[illegible] |
| Unknown | 1 | 0 | | | 0 | 0 | 1 | | |
| Died in hospital | 181 (7.0%) | 8 (29%) | <0.001 | **0.004** | 4 (7.8%) | 8 (12%) | 177 (7.1%) | 0.2 | 0.3 |
| Number of diagnostics | 13 (9, 19) | 17 (10, 27) | 0.009 | **0.028** | 16 (13, [illegible] | 17 (9, 24) | 13 (9, 19) | <0.001 | **<0.00**[illegible] |

### eTable 10. Univariate comparisons between clusters of RDW and Red Blood Cells

| | RDW | | | | | Red Blood Cells | | | | |
|---|---|---|---|---|---|---|---|---|---|---|
| **Characteristic** | **1**<br>**N = 234** | **2**<br>**N = 2267** | **3**<br>**N = 102** | **p valu e** | **q valu e** | **1**<br>**N = 493** | **2**<br>**N = 285** | **3**<br>**N = 1825** | **p value** | **q value** |
| Age | 71 (56, 83) | 62 (42, 79) | 56 (37, 79) | <0.00 | **<0.00** | 62 (40, | 43 (29, 59) | 65 (47, 81) | <0.00 | **<0.00** |
| Gender | | | | 0.015 | **0.026** | | | | <0.00 | **<0.00** |
| F | 115 (49%) | 897 (40%) | 43 (42%) | | | 165 | 61 (21%) | 829 (45%) | | |
| M | 119 (51%) | 1,370 | 59 (58%) | | | 328 | 224 (79%) | 996 (55%) | | |
| Ethnicity | | | | 0.12 | 0.14 | | | | 0.042 | **0.042** |
| ASIAN | 8 (3.7%) | 39 (2.0%) | 2 (2.7%) | | | 10 (2.2%) | 6 (2.7%) | 33 (2.1%) | | |
| BLACK | 20 (9.3%) | 103 (5.2%) | 2 (2.7%) | | | 31 (6.9%) | 6 (2.7%) | 88 (5.5%) | | |
| HISPANIC | 8 (3.7%) | 87 (4.4%) | 3 (4.0%) | | | 26 (5.8%) | 15 (6.7%) | 57 (3.6%) | | |
| MULTI | 0 (0%) | 5 (0.3%) | 1 (1.3%) | | | 1 (0.2%) | 1 (0.4%) | 4 (0.3%) | | |
| OTHER | 6 (2.8%) | 96 (4.9%) | 4 (5.3%) | | | 22 (4.9%) | 16 (7.2%) | 68 (4.3%) | | |
| WHITE | 172 (80%) | 1,637 | 63 (84%) | | | 357 | 179 (80%) | 1,336 | | |
| Unknown | 20 | 300 | 27 | | | 46 | 62 | 239 | | |
| Cohort | | | | 0.089 | 0.13 | | | | <0.00 | **<0.00** |
| SCI | 25 (11%) | 334 (15%) | 23 (23%) | | | 45 (9.1%) | 66 (23%) | 271 (15%) | | |
| SCI noFracture | 10 (4.3%) | 112 (4.9%) | 3 (2.9%) | | | 40 (8.1%) | 9 (3.2%) | 76 (4.2%) | | |
| Spine | 199 (85%) | 1,821 | 76 (75%) | | | 408 | 210 (74%) | 1,478 | | |
| Length of stay | 8 (5, 13) | 7 (4, 12) | 9 (6, 11) | 0.2 | 0.2 | 5 (4, 9) | 8 (5, 16) | 8 (5, 13) | <0.00 | **<0.00** |
| Unknown | 0 | 1 | 0 | | | 0 | 0 | 1 | | |
| Died in hospital | 31 (13%) | 140 (6.2%) | 18 (18%) | <0.00 | **0.001** | 20 (4.1%) | 25 (8.8%) | 144 | 0.005 | **0.006** |
| Number of | 20 (14, 25) | 13 (9, 19) | 15 (10, 23) | <0.00 | **<0.00** | 12 (8, 17) | 13 (9, 20) | 14 (9, 20) | <0.00 | **<0.00** |

### eTable 11. Univariate comparisons between clusters of White blood cells

| Characteristic | White Blood Cells 1 N = 22 | 2 N = 2553 | 3 N = 28 | p value | q value* |
|---|---|---|---|---|---|
| Age | 82 (65, 88) | 62 (43, 79) | 69 (58, 82) | 0.007 | **0.015** |
| Gender | | | | 0.7 | 0.9 |
| F | 7 (32%) | 1,037 (41%) | 11 (39%) | | |
| M | 15 (68%) | 1,516 (59%) | 17 (61%) | | |
| Ethnicity | | | | >0.9 | >0.9 |
| ASIAN | 0 (0%) | 49 (2.2%) | 0 (0%) | | |
| BLACK | 1 (5.9%) | 123 (5.6%) | 1 (4.0%) | | |
| HISPANIC | 0 (0%) | 98 (4.4%) | 0 (0%) | | |
| MULTI RACE | 0 (0%) | 6 (0.3%) | 0 (0%) | | |
| OTHER | 1 (5.9%) | 104 (4.7%) | 1 (4.0%) | | |
| WHITE | 15 (88%) | 1,834 (83%) | 23 (92%) | | |
| Unknown | 5 | 339 | 3 | | |
| Cohort | | | | 0.015 | **0.026** |
| SCI Fracture | 5 (23%) | 368 (14%) | 9 (32%) | | |
| SCI noFracture | 0 (0%) | 122 (4.8%) | 3 (11%) | | |
| Spine Trauma | 17 (77%) | 2,063 (81%) | 16 (57%) | | |
| Length of stay (days) | 7 (3, 13) | 7 (5, 13) | 7 (4, 9) | 0.3 | 0.4 |
| Unknown | 0 | 1 | 0 | | |
| Died in hospital | 8 (36%) | 172 (6.7%) | 9 (32%) | <0.001 | **0.002** |
| Number of diagnostics | 26 (17, 33) | 13 (9, 19) | 15 (11, 21) | <0.001 | **<0.001** |

### eFigures

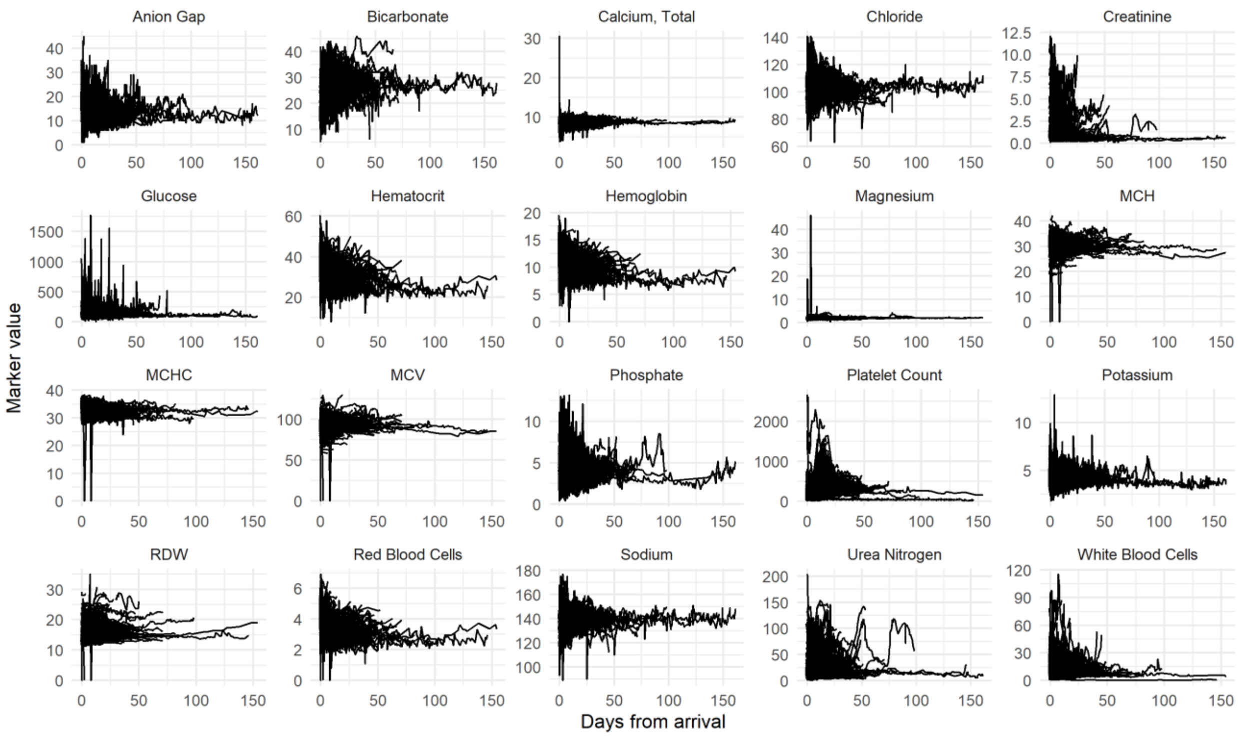


**eFigure 1. Spaghetti plots for the raw data of the modeling set of analytes (20 most common).** Note that unexpected spikes are observed, probably indicative of data errors. Each line represents a subject. As expected by the nature of the data, time is asynchronous, meaning that markers were obtained for each patient at different timepoints in non-regular intervals. No apparent trends are observable from the plots. We can observe fluctuations over time, with, in general, a high dynamic range early after admission that reduces as time progresses.

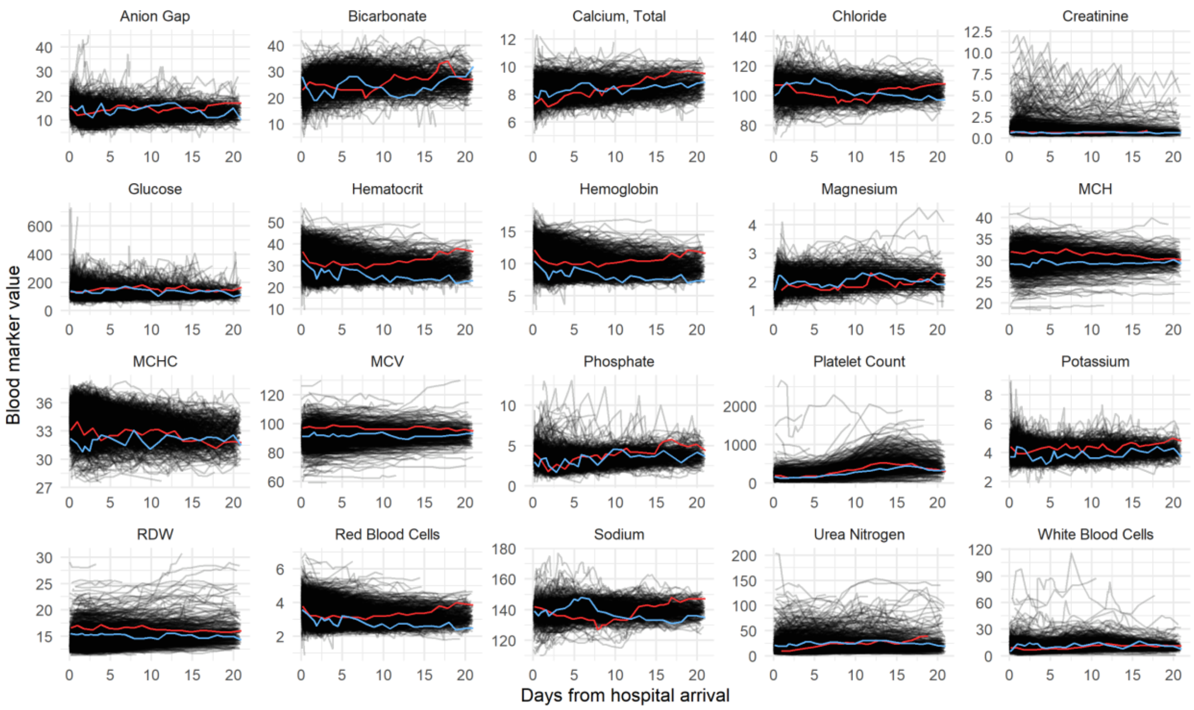


**eFigure 2. Spaghetti plots for the outlier-cleaned modeling set of laboratory analytes for the first 21 days after admission.** Each line represents a single subject. Lines red and blue are two randomly selected subjects illustrating differences in temporal trends.

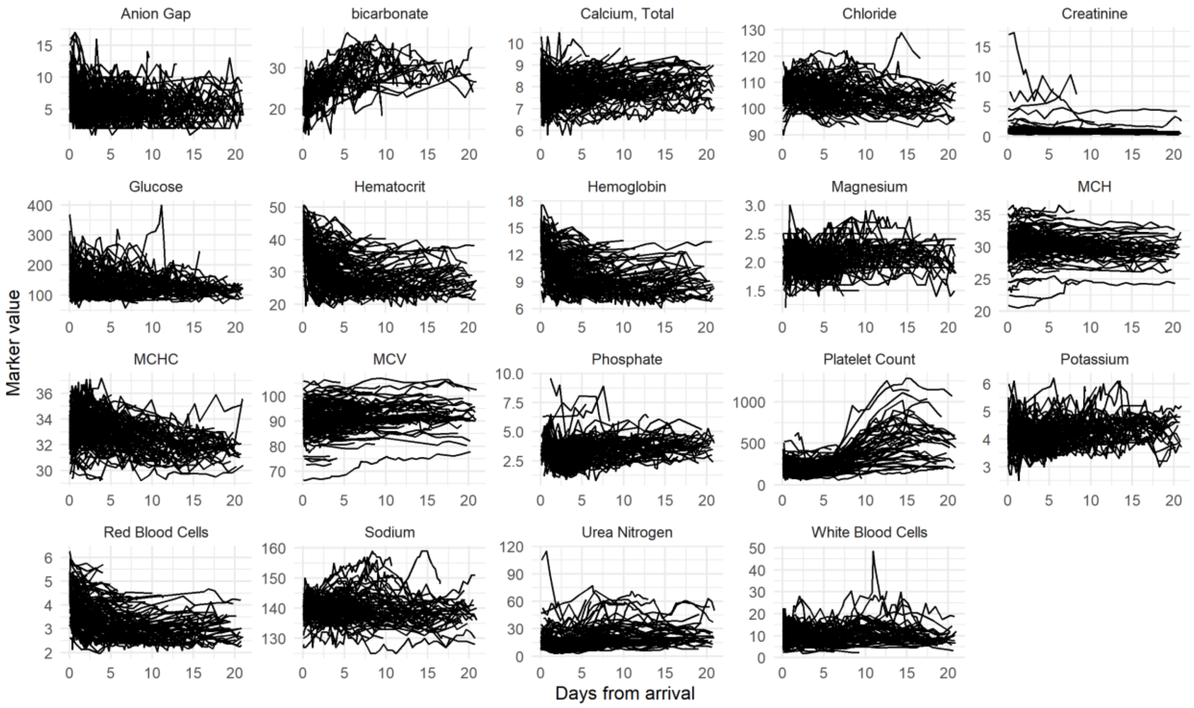

**eFigure 3. Spaghetti plots for the outlier-cleaned TRACK-SCI minimal set of laboratory analytes for the first 21 days after admission.** Each line represents a single subject.

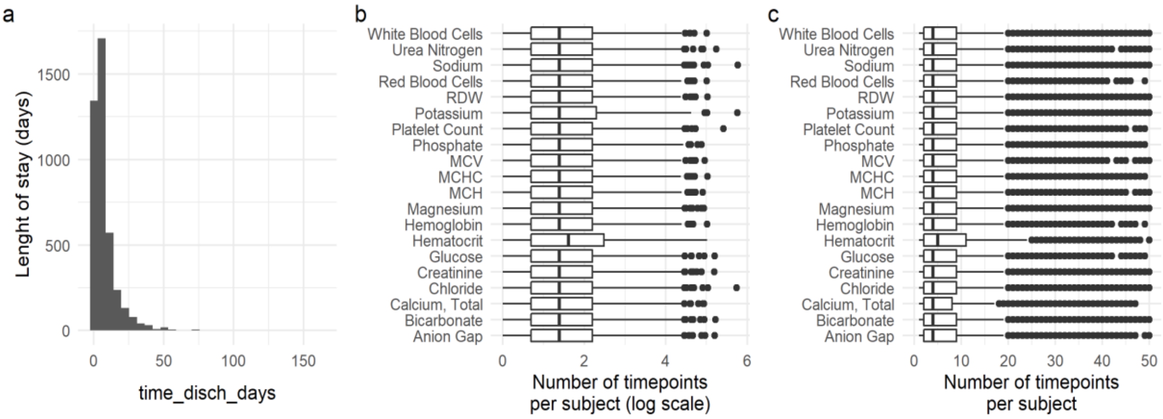


**eFigure 4. Length of stay distribution**. (a) Histogram of the length of stay in days for the cohort. (b) Logarithmic scale count of the number of measurements per subject and the modeling set of analytes. (c) Count of the number of measurements per subject and the modeling set of analytes. The number of subjects with data in a given marker also reduces over time, with very few subjects with data beyond 50 days (eFigure 1). This can also be confirmed by the distribution of length of stay, where the median time is 4.6 days (first quartile: 2.09 and third quartile: 9.07), and 96.4% of the cohort was discharged before 18 days in the hospital.

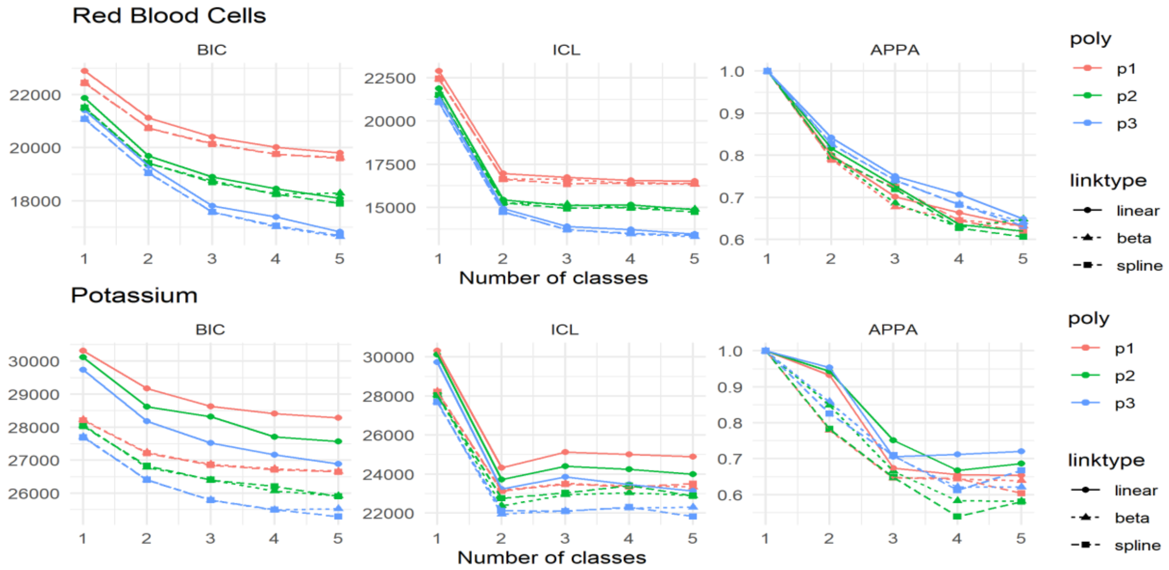


**eFigure 5. Example of model fit plots for two different types of model selection "patterns".** Top row shows the BIC, ICL and APPA (mean APPA across classes) for Red Blood Cells. It can be observed that polynomial degree has a higher effect on BIC and ICL than the type of link function. For Potassium (bottom row), the effect of polynomial degree and type of link function is

compounded. The higher drop in ICL for both markers from 1 to 2 classes suggests that the major gain in model fit happens when more than 1 class is considered, illustrating the need for modeling heterogeneous populations.

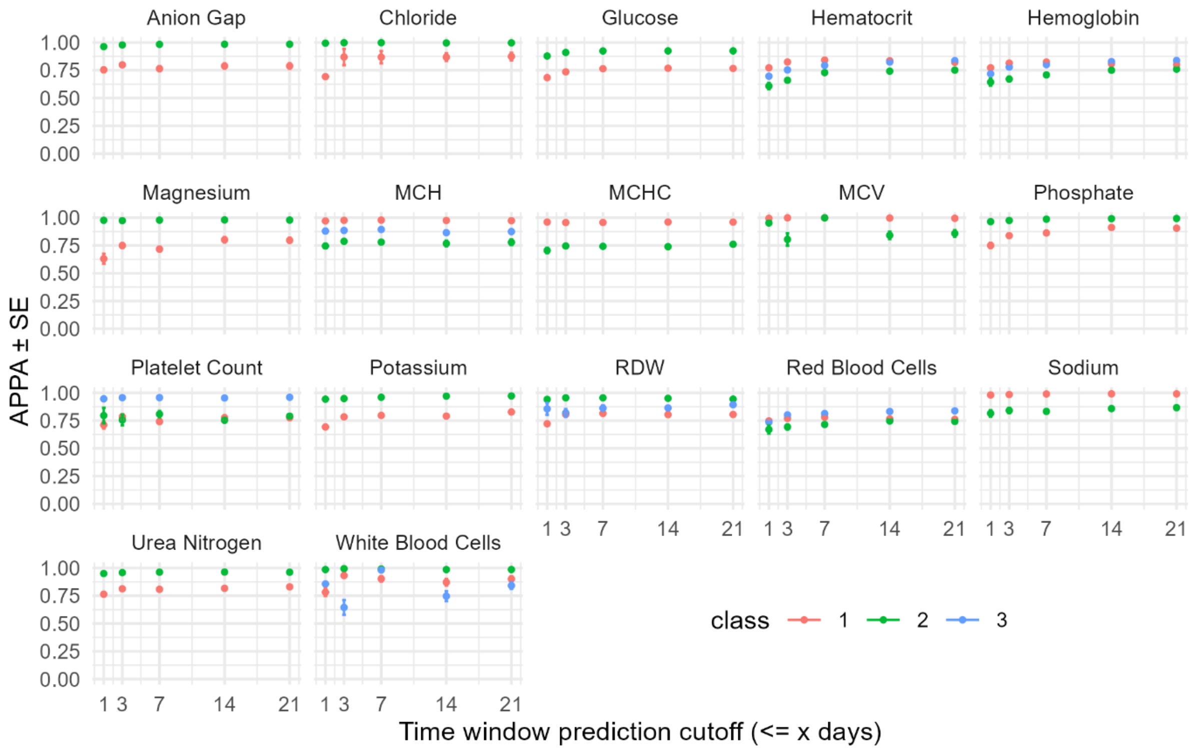


**eFigure 6. Average posterior probability of assignment (APPA) for each trajectory class and marker at each time point window in MIMIC.** Each point represents the APPA of using the respective model and <= x days of data to predict trajectory membership, where x is the time window cutoff. The standard error of the mean (SE) across PPA for all subjects is represented. Markers with a single trajectory class are not shown as they have an APPA = 1.

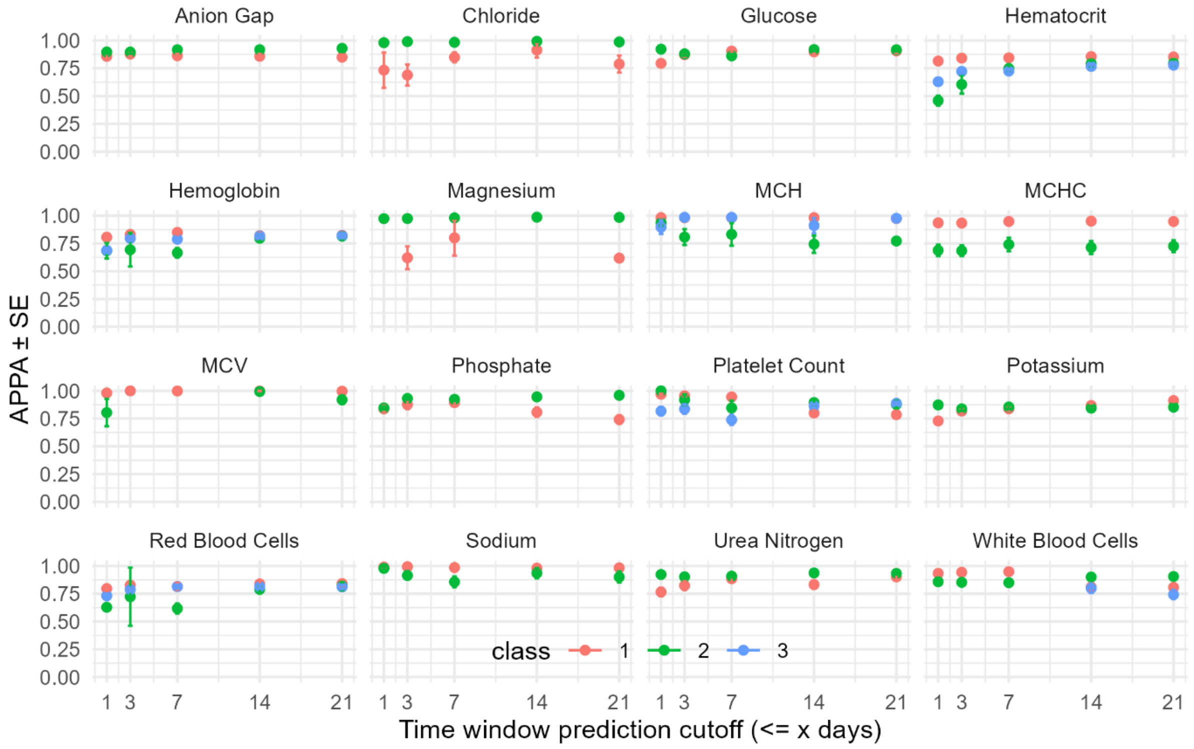


**eFigure 7. Average posterior probability of assignment (APPA) for each trajectory class and marker at each time point window in TRACK_SCI.** Each point represents the APPA of using the respective model and <= x days of data to predict trajectory membership, where x is the time window cutoff. The standard error of the mean (SE) across PPA for all subjects is represented. Markers with a single trajectory class are not shown as they have an APPA = 1.

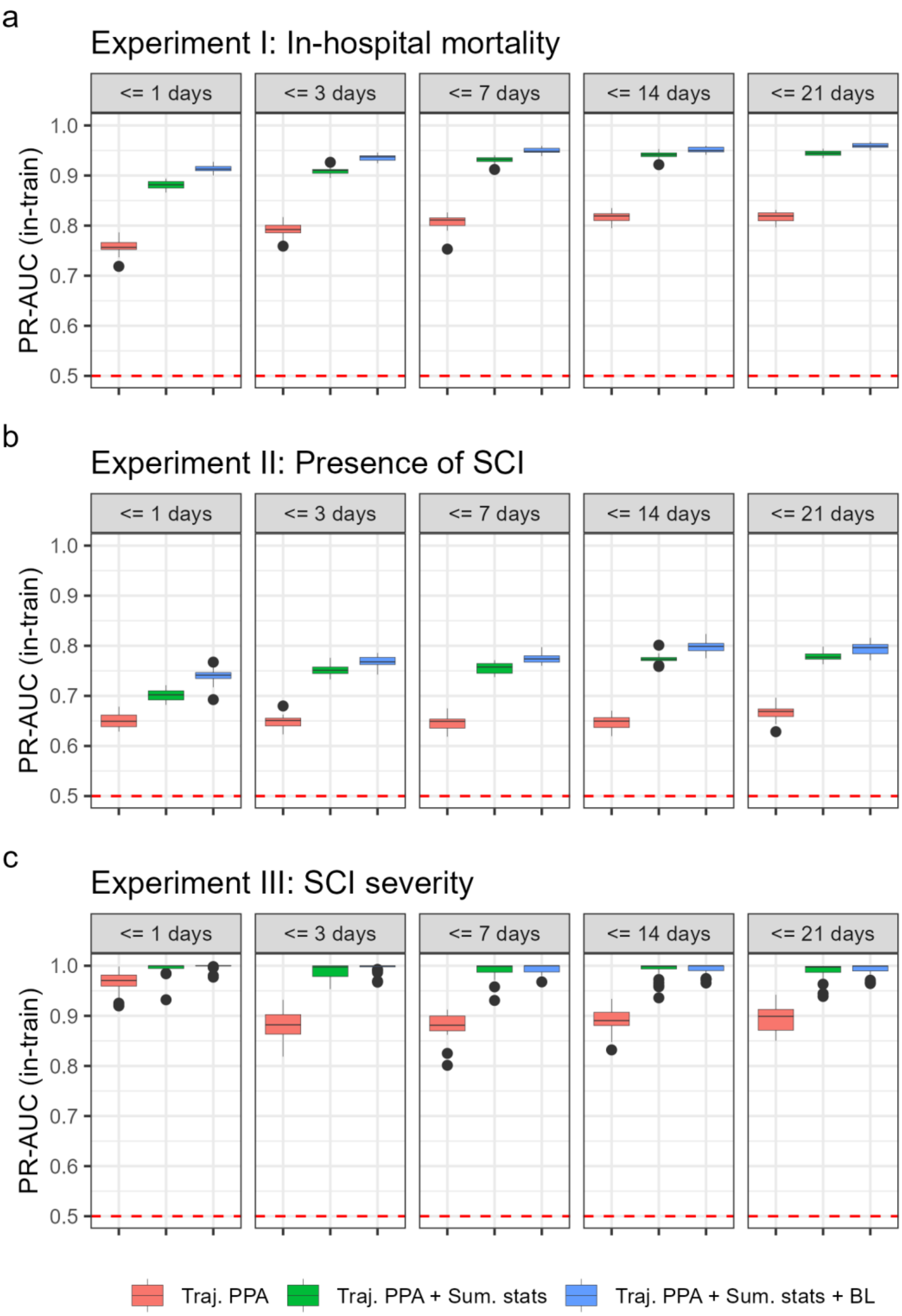


**eFigure 8. PR-AUC performance of dynamic predictions on the train data.** (a) PR-AUC in-train sample performance of task I in-hospital mortality. (b) PR-AUC in-train sample performance of task II for detecting the presence of SCI after spine trauma. (c) PR-AUC in-train sample performance of task III on detecting SCI severity on the TRACK-SCI cohort, external to trajectory modeling. Dashed red lines represent the mean prevalence of the outcome of interest in each experiment. Three predictors' lists are shown: Traj. PPA = posterior probability of trajectory classification only; + Sum. stats = addition of summary statistics of blood data; and + BL = addition of baseline predictors.

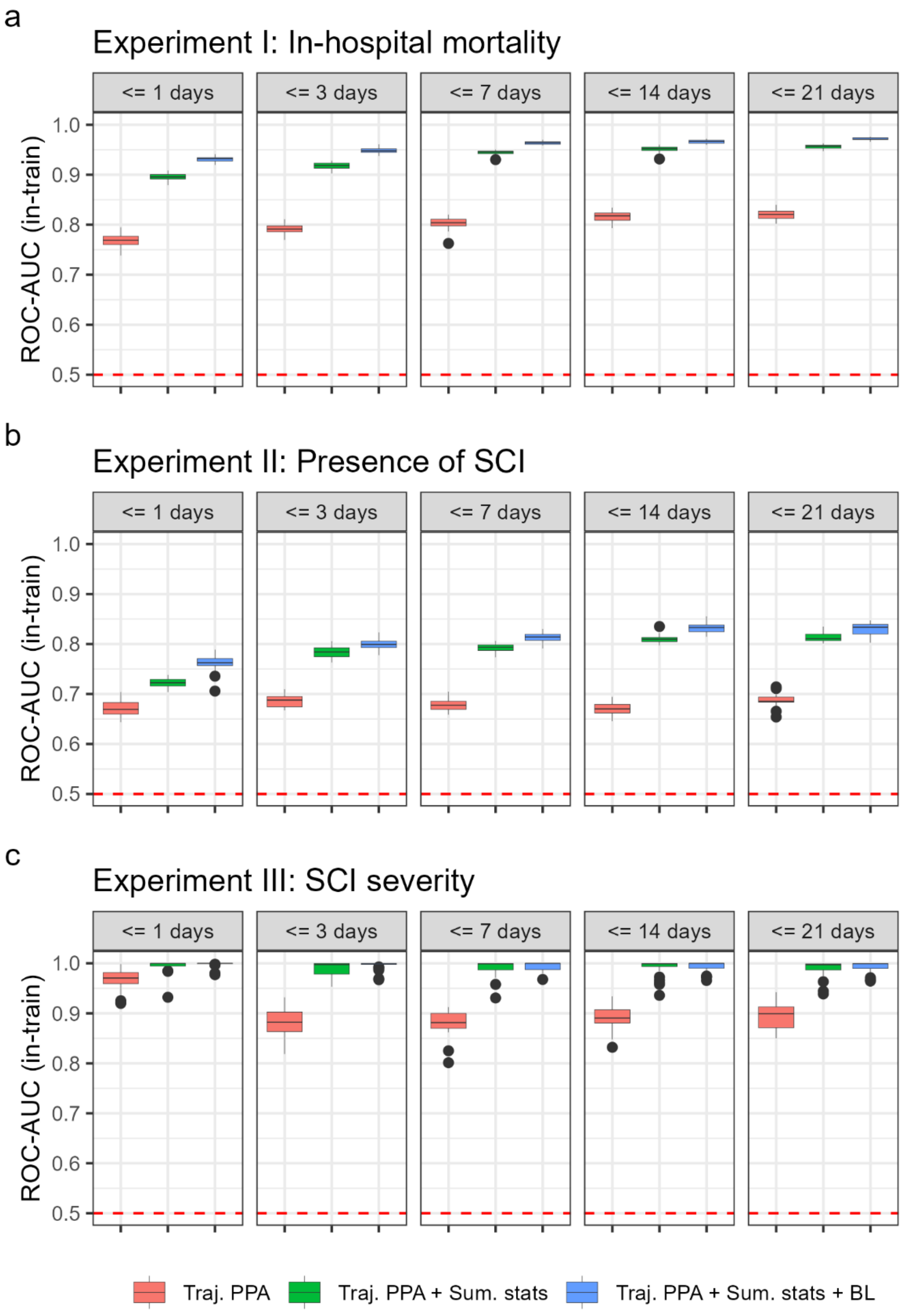


**eFigure 9. ROC-AUC performance of dynamic predictions on the train data.** (a) ROC-AUC in-train sample performance of task I in-hospital mortality. (b) ROC-AUC in-train sample performance of task II for detecting the presence of SCI after spine trauma. (c) ROC-AUC in-train sample performance of task III on detecting SCI severity on the TRACK-SCI cohort, external to trajectory modeling. Dashed red lines represent the mean prevalence of the outcome of interest in each experiment. Three predictors' lists are shown: Traj. PPA = posterior probability of trajectory classification only; + Sum. stats = addition of summary statistics of blood data; and + BL = addition of baseline predictors.

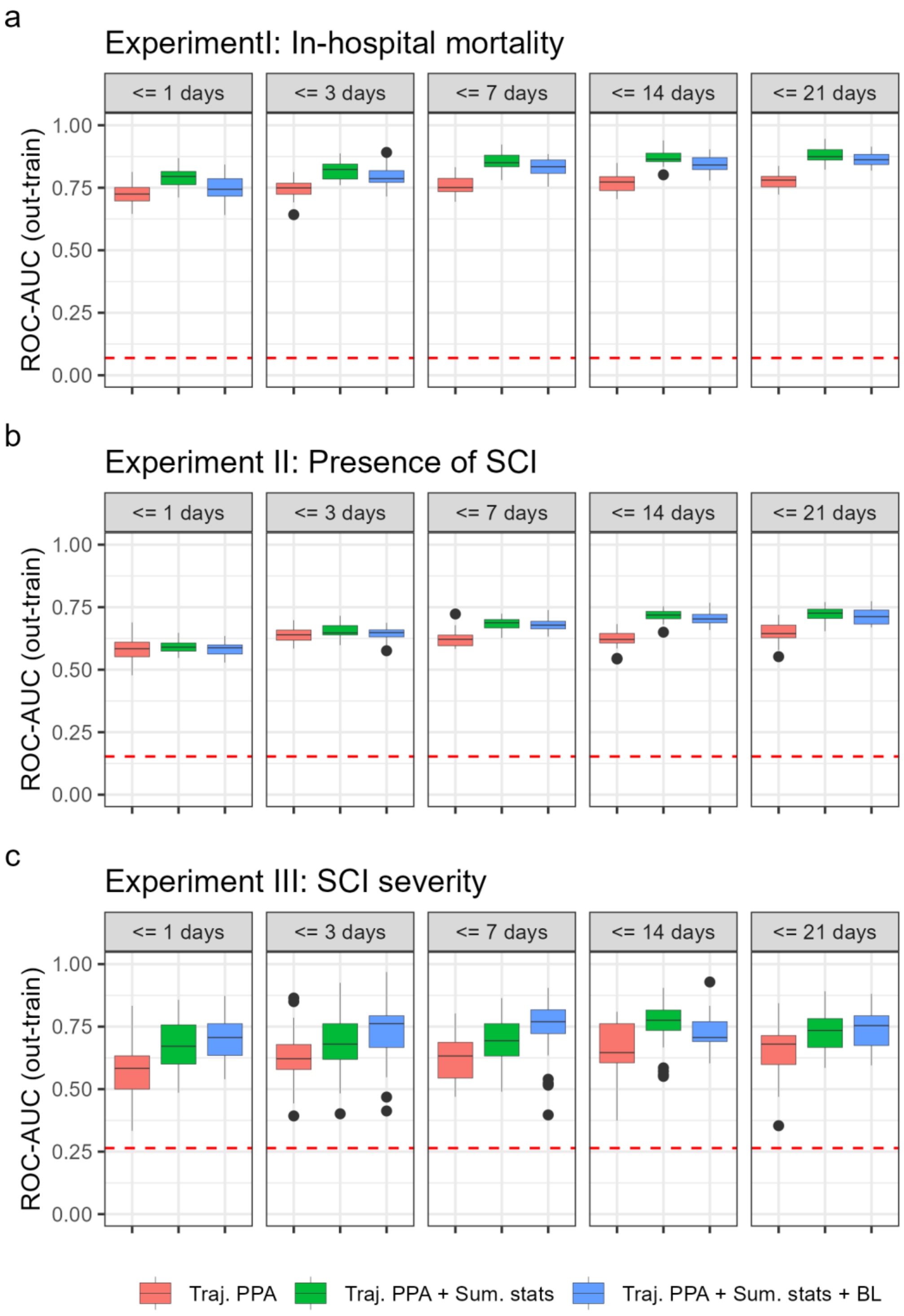


**eFigure 10. ROC-AUC performance of dynamic predictions on the out-train data.** (a) ROC-AUC out-train sample performance of task I in-hospital mortality. (b) ROC-AUC out-train sample performance of task II for detecting the presence of SCI after spine trauma. (c) ROC-AUC out-train sample performance of task III on detecting SCI severity on the TRACK-SCI cohort, external to trajectory modeling. Dashed red lines represent the mean prevalence of the outcome of interest in each experiment. Three predictors' lists are shown: Traj. PPA = posterior probability of trajectory classification only; + Sum. stats = addition of summary statistics of blood data; and + BL = addition of baseline predictors.

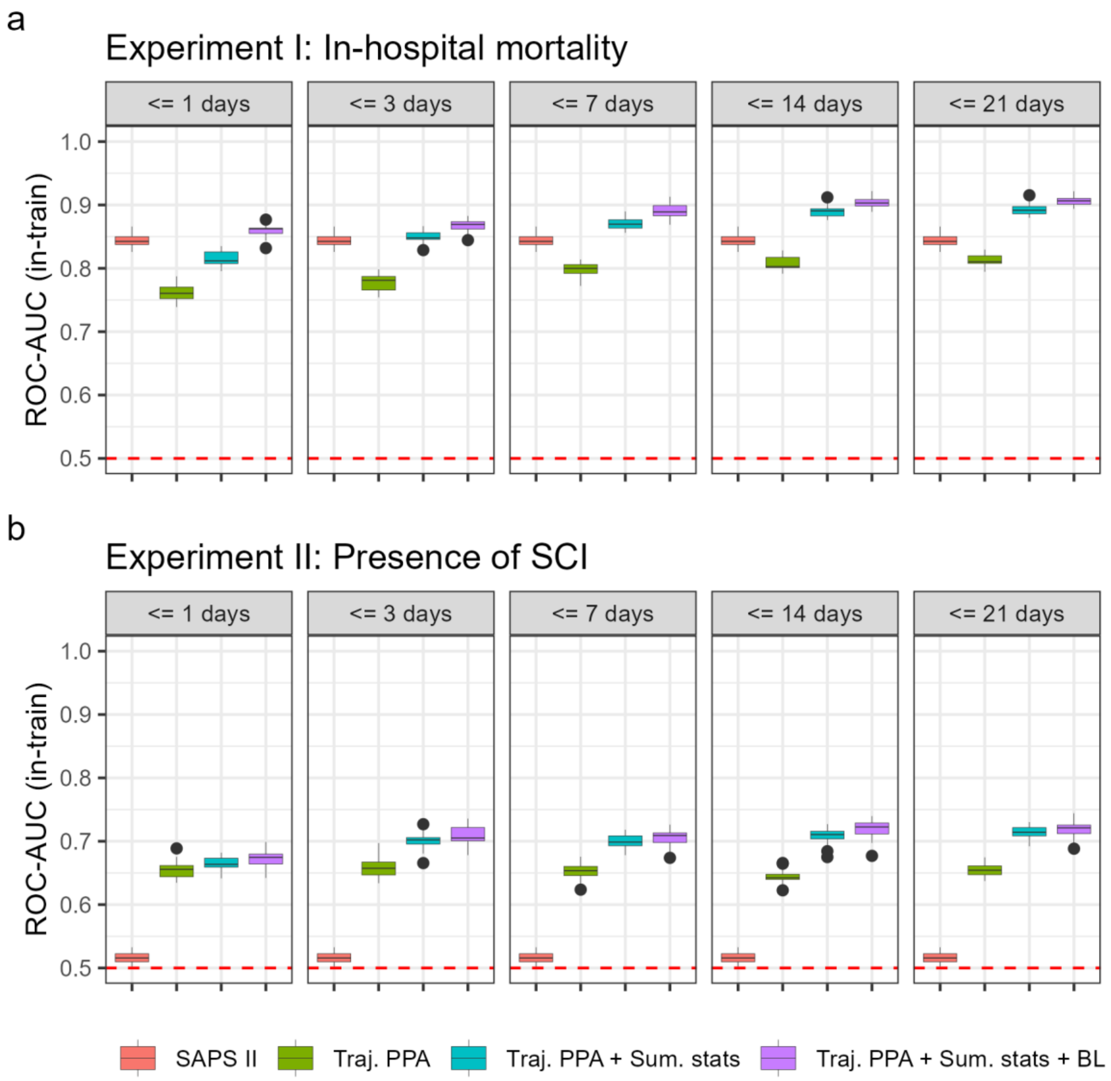


**eFigure 11. ROC-AUC performance of dynamic predictions on the train data for ICU patients.** (a) ROC-AUC in-train sample performance of task I in-hospital mortality. (b) ROC-AUC in-train sample performance of task II for detecting the presence of SCI after spine trauma. Dashed red lines represent the mean prevalence of the outcome of interest in each experiment. Four predictors' lists are shown: SAPS = SAPS II score; Traj. PPA = posterior probability of trajectory classification only; + Sum. stats = addition of summary statistics of blood data; and + BL = addition of baseline predictors.

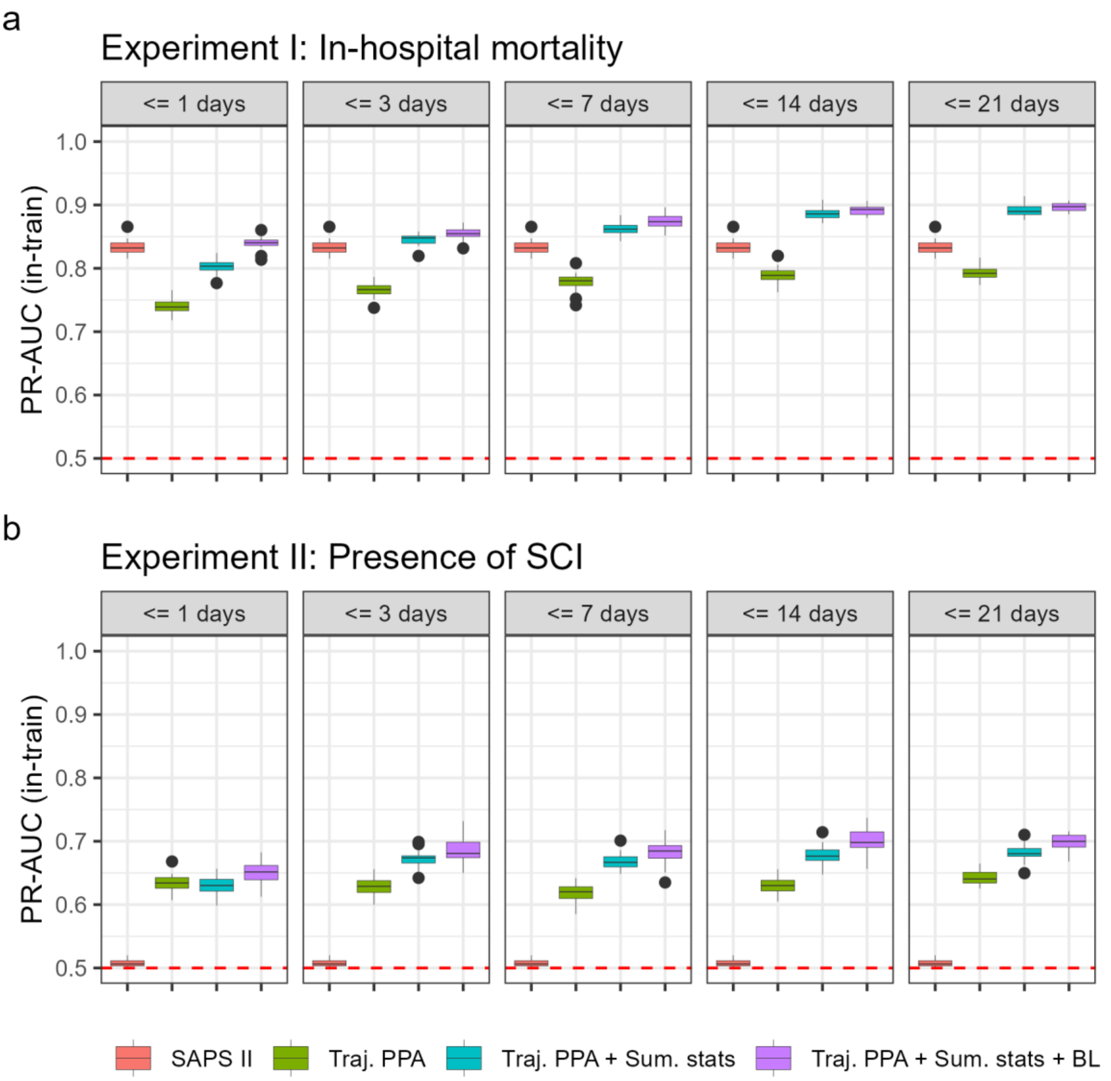


**eFigure 12. PR-AUC performance of dynamic predictions on the train data for ICU patients.** (a) PR-AUC in-train sample performance of task I in-hospital mortality. (b) PR-AUC in-train sample performance of task II for detecting the presence of SCI after spine trauma. Dashed red lines represent the mean prevalence of the outcome of interest in each experiment. Four predictors' lists are shown: SAPS = SAPS II score; Traj. PPA = posterior probability of trajectory classification only; + Sum. stats = addition of summary statistics of blood data; and + BL = addition of baseline predictors.

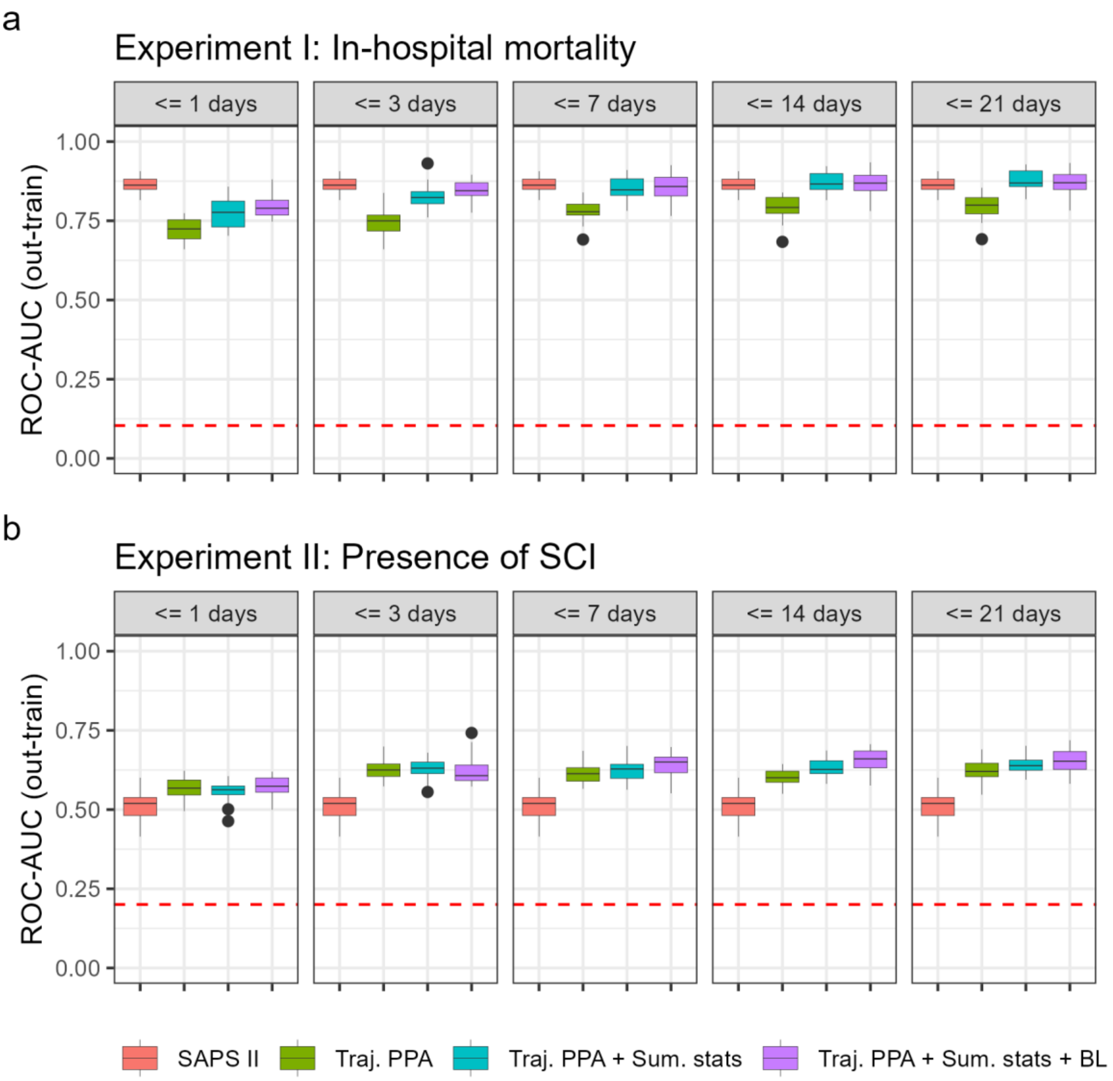


**eFigure 13. ROC-AUC performance of dynamic predictions on the out-train data for ICU patients.** (a) ROC-AUC out-train sample performance of task I in-hospital mortality. (b) ROC-AUC out-train sample performance of task II for detecting the presence of SCI after spine trauma. Dashed red lines represent the mean prevalence of the outcome of interest in each experiment. Four predictors' lists are shown: SAPS = SAPS II score; Traj. PPA = posterior probability of trajectory classification only; + Sum. stats = addition of summary statistics of blood data; and + BL = addition of baseline predictors.

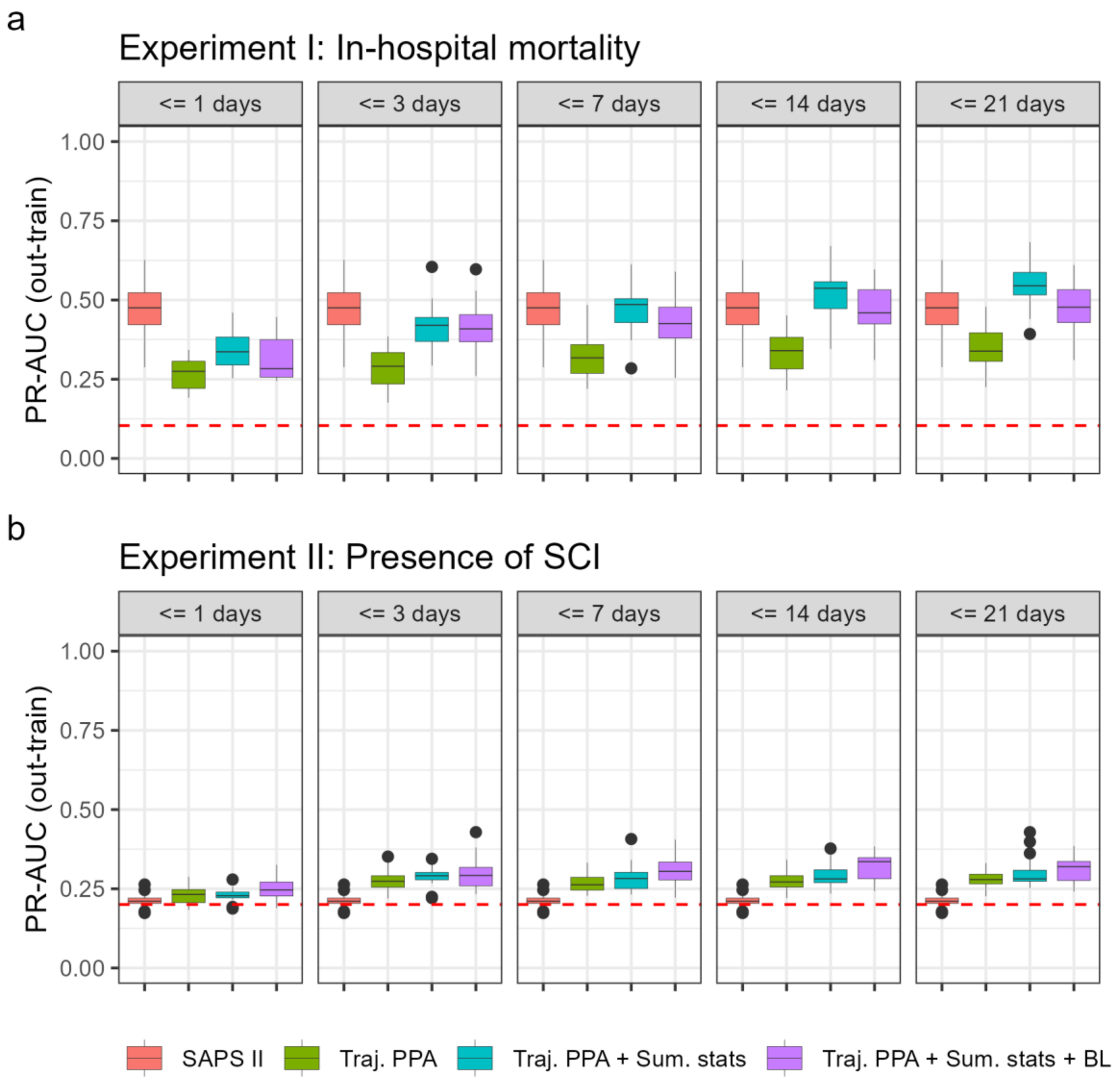


**eFigure 14. PR-AUC performance of dynamic predictions on the out-train data for ICU patients.** (a) PR-AUC out-train sample performance of task I in-hospital mortality. (b) PR-AUC out-train sample performance of task II for detecting the presence of SCI after spine trauma. Dashed red lines represent the mean prevalence of the outcome of interest in each experiment. Four predictors' lists are shown: SAPS = SAPS II score; Traj. PPA = posterior probability of trajectory classification only; + Sum. stats = addition of summary statistics of blood data; and + BL = addition of baseline predictors.

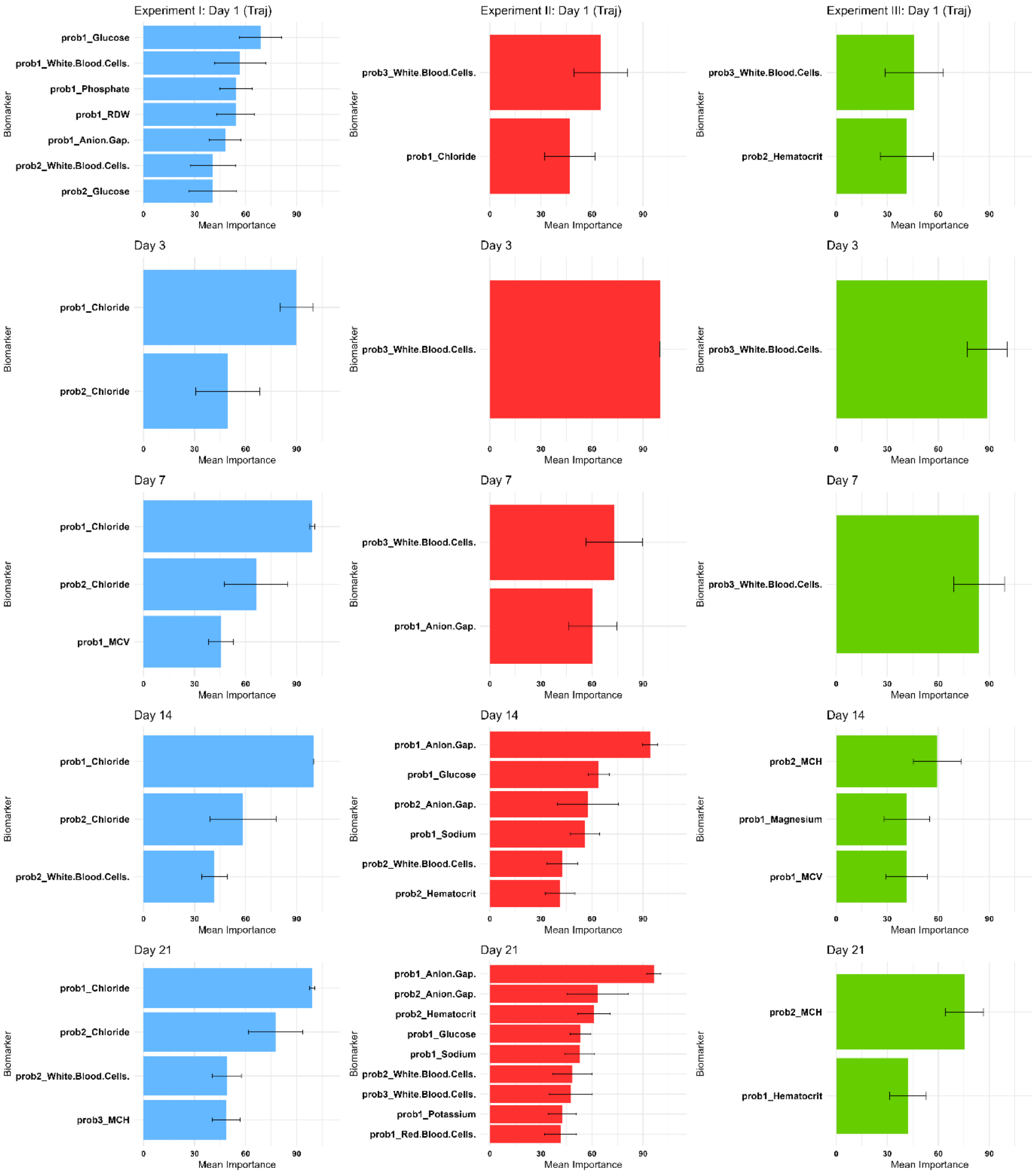


**eFigure 15. Variable importance for models with posterior probability of trajectory classification only.** Variables with mean importance of more than 40 for (Left) Experiment I: In-hospital mortality; (Middle) Experiment II: presence of SC; (Right) Experiment III: severity of SCI.

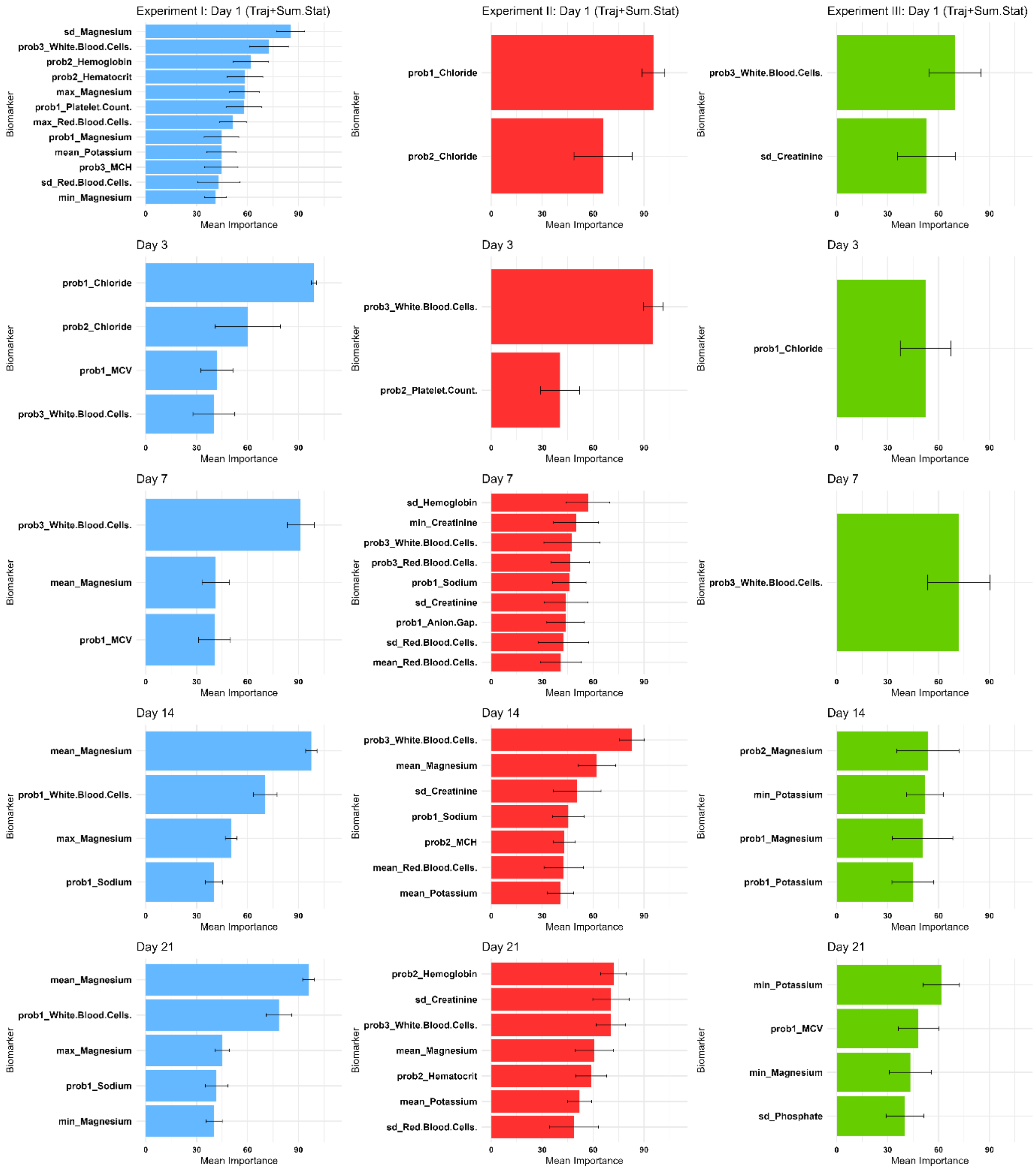


**eFigure 16. Variable importance for models with posterior probability of trajectory classification and summary statistics of biomarkers.** Variables with mean importance of more than 40 for (Left) Experiment I: In-hospital mortality; (Middle) Experiment II: presence of SC; (Right) Experiment III: severity of SCI.

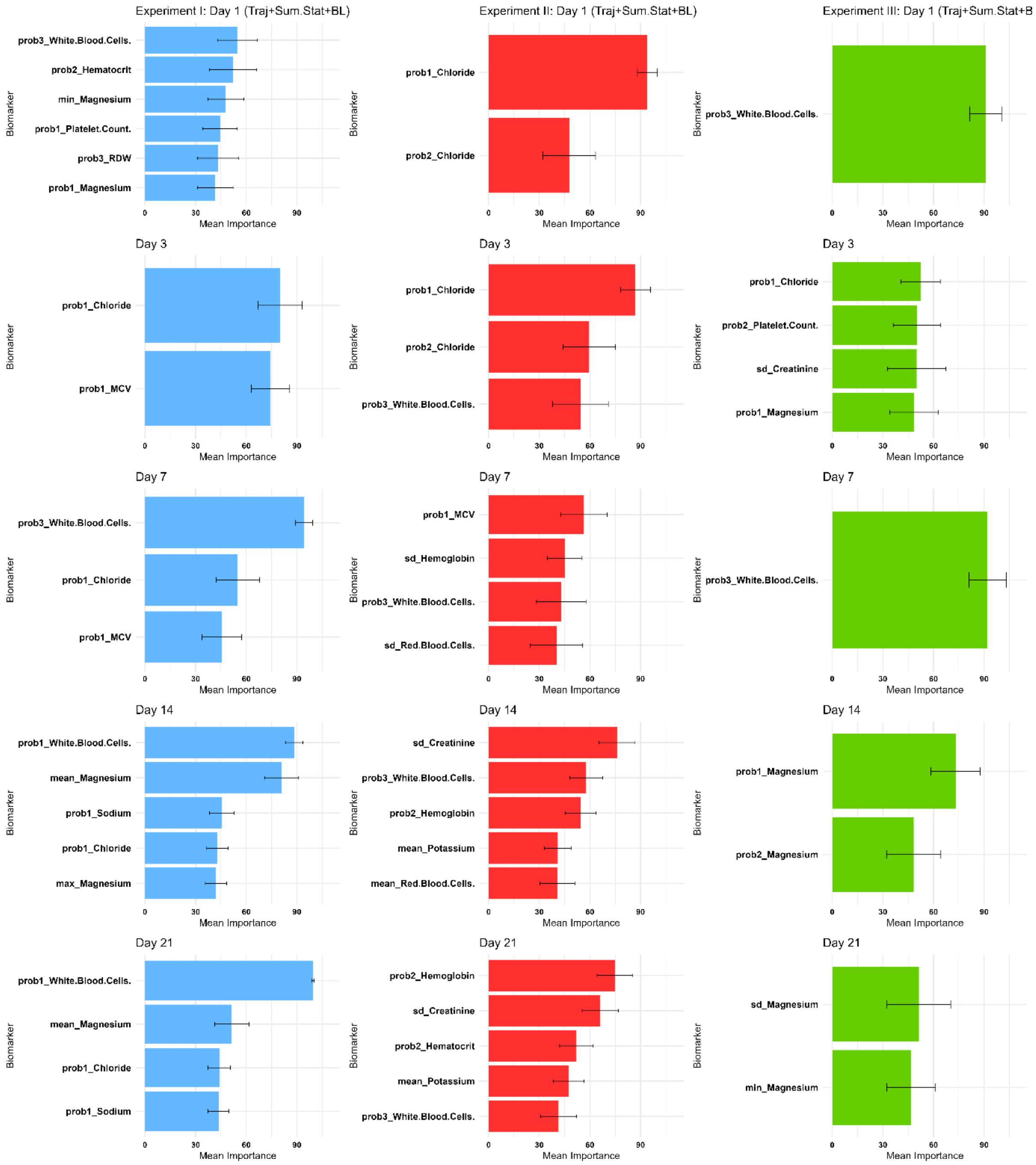


**eFigure 17. Variable importance for models with posterior probability of trajectory classification, summary statistics and baseline predictors.** Variables with mean importance of more than 40 for (Left) Experiment I: In-hospital mortality; (Middle) Experiment II: presence of SC; (Right) Experiment III: severity of SCI.

**References**


1. Tukey JW. *Exploratory Data Analysis*. Addison-Wesley Pub. Co.; 1977.

2. Seo S. A Review and Comparison of Methods for Detecting Outliers in Univariate Data Sets. *undefined*. Published online 2006. Accessed March 8, 2022. https://www.semanticscholar.org/paper/A-Review-and-Comparison-of-Methods-for-Detecting-in-Seo/cb868f0b242b9623b7544a58b6a21647dfa138a5

3. Le Gall JR, Lemeshow S, Saulnier F. A new simplified acute physiology score (SAPS II) based on a European/North American multicenter study. *Jama*. 1993;270(24):2957-2963.
